# Migrants’ primary care utilisation before and during the COVID-19 pandemic in England: An interrupted time series

**DOI:** 10.1101/2022.03.14.22272283

**Authors:** Claire X. Zhang, Yamina Boukari, Neha Pathak, Rohini Mathur, Srinivasa Vittal Katikireddi, Parth Patel, Ines Campos-Matos, Dan Lewer, Vincent Nguyen, Greg Hugenholtz, Rachel Burns, Amy Mulick, Alasdair Henderson, Robert W. Aldridge

## Abstract

**Background:** How international migrants access and use primary care in England is poorly understood. We aimed to compare primary care consultation rates between international migrants and non-migrants in England before and during the COVID-19 pandemic (2015– 2020).

**Methods:** Using linked data from the Clinical Practice Research Datalink (CPRD) GOLD and the Office for National Statistics, we identified migrants using country-of-birth, visa-status or other codes indicating international migration. We ran a controlled interrupted time series (ITS) using negative binomial regression to compare rates before and during the pandemic.

**Findings:** In 262,644 individuals, pre-pandemic consultation rates per person-year were 4.35 (4.34-4.36) for migrants and 4.6 (4.59-4.6) for non-migrants (RR:0.94 [0.92-0.96]). Between 29 March and 26 December 2020, rates reduced to 3.54 (3.52-3.57) for migrants and 4.2 (4.17-4.23) for non-migrants (RR:0.84 [0.8–0.88]). Overall, this represents an 11% widening of the pre-pandemic difference in consultation rates between migrants and non-migrants during the first year of the pandemic (RR:0.89, 95%CI:0.84–0.94). This widening was greater for children, individuals whose first language was not English, and individuals of White British, White non-British and Black/African/Caribbean/Black British ethnicities.

**Interpretation:** Migrants were less likely to use primary care before the pandemic and the first year of the pandemic exacerbated this difference. As GP practices retain remote and hybrid models of service delivery, they must improve services and ensure they are accessible and responsive to migrants’ healthcare needs.

**Funding:** This study was funded by the Medical Research Council (MR/V028375/1) and Wellcome Clinical Research Career Development Fellowship (206602).

## Background

Effective primary care is linked to better health outcomes in the general population (1). In the United Kingdom (UK), access to primary care is free of charge for all (2). However, barriers to general practice (GP) registration and low registration rates are long-standing issues among international migrants (3-7). For migrants who do register, there are barriers to accessing care, including insufficient translation support, discrimination, and transportation costs (8-10). However, differences in primary care utilisation between migrants and the UK-born population are poorly understood and have relied on self-reported surveys with limited sample sizes and mixed results (11, 12). Examining these differences is of particular importance for service planning given the UK is home to the fifth largest number of international migrants in the world (13).

Substantial reductions in primary care consultations were observed across the UK during the first wave of the COVID-19 pandemic (14, 15), with people from minority ethnic groups reporting greater healthcare disruption than individuals from White ethnic groups (16). However, the difference in primary care utilisation between migrants and non-migrants in England and how the pandemic has affected this, including the interplay between migration and ethnicity, has not been studied. This is needed to help identify inequalities, and to inform service provision and policy (17). Shifts from in-person to remote consultations may make primary care access even more challenging for people at risk of digital exclusion, including some migrants (18), although no association was found between deprivation status and the likelihood of accessing remote consultations (19).

Recent validation of an electronic health record (EHR) code list to identify a cohort of migrants largely representative of the broader migrant population (20) presents a unique opportunity to quantify differences in primary care usage between migrants and non-migrants in England before and during the COVID-19 pandemic.

This study aimed to compare National Health Service (NHS) primary care consultation rates between migrants and non-migrants from 2015 to 2020 in England, specifically:

1. Did consultation rates differ between migrants and non-migrants before the pandemic?
2. Did this change during the pandemic?
3. Did differences between migrants and non-migrants vary across ethnic groups?

## Methods

### Study design and data management

Of over 900 GP practices in the UK contributing to Clinical Practice Research Datalink (CPRD) GOLD, 413 were in England and had linked Office of National Statistics (ONS) 2015 Index of Multiple Deprivation (IMD) data. Data flows are shown in Figure 1. Pseudo-anonymised data were stored, cleaned and analysed using R (versions 3.6.2 and 4.0.3) in the University College London Data Safe Haven during 2021. All code for data cleaning and analysis is freely available.

**Figure 1:**
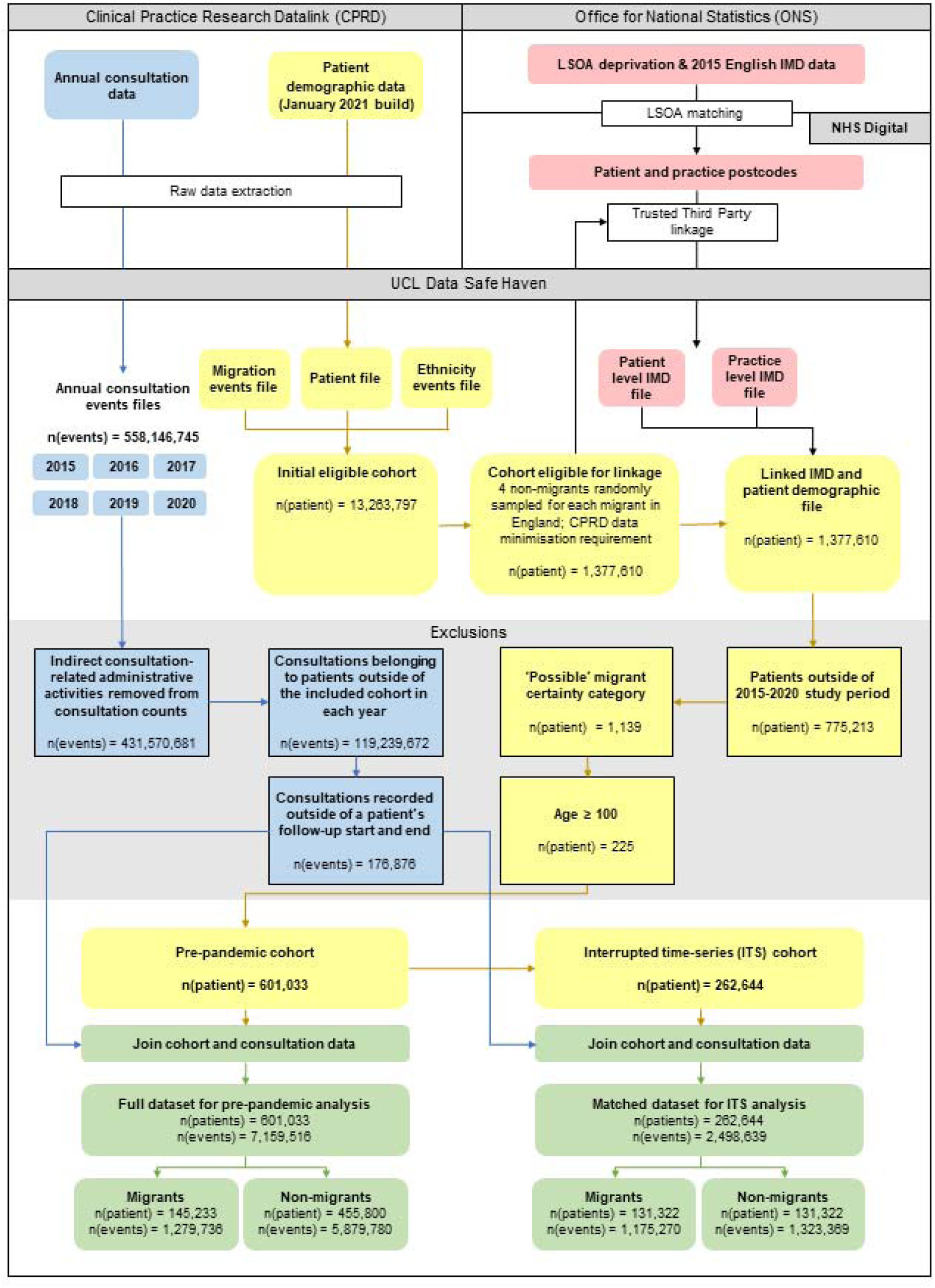
Data flow diagram with patient and consultation exclusions

### Exposure and outcomes

The exposure of interest was international migration to the UK, determined by a validated migration code list comprising migration-related ‘Read Version 2’ codes (20). The code list produces a binary indicator (migrants, non-migrants). It also disaggregates migrants into levels of certainty: ‘definite’ migrants with country-of-birth and/or visa-status codes, ‘probable’ migrants with main-language-other-than-English codes, and ‘possible’ migrants with non-UK-origin codes.

The outcomes were primary care consultation rates (per person-year) and rate ratios (RRs) comparing migrants to non-migrants. We derived consultation counts using 28 out of 62 consultation types in CPRD indicating direct consultations (as opposed to indirect administrative activities) and further disaggregated them into face-to-face and telephone consultations; the latter of which may also include other types of virtual consultations (Table S1).

### Study cohort

The initial eligible cohort comprised individuals of all ages registered before January 2021, for any length of time, at a CPRD GOLD GP practice in the UK that was contributing ‘up-to-standard’ data (see Supplementary Box 1 for details) for any length of time in the January 2021 database build (21). We reduced this initial cohort to individuals registered at a GP practice in England who were eligible for all linkages pre-specified in the study protocol (22) and we identified migrants by applying the migration code list. To comply with CPRD’s data minimisation policy, we randomly sampled non-migrants from the reduced initial cohort at a ratio of 1:4 migrants to non-migrants and then linked this cohort to IMD data.

Follow-up commenced at the latest date of a) an individual registering at a CPRD GOLD practice; b) their practice’s first ‘up-to-standard’ date; c) 1 January 2015. Follow-up ended at the earliest date of a) a patient’s transfer out of a CPRD GOLD practice; b) their date of death; c) the last data collection date for the practice; d) 26 December 2020 (end date of available data).

We made exclusions at the patient and consultation level (Figure 1 and Supplementary Box 1). To prioritise specificity, we excluded ‘possible’ migrants due to uncertainty around their migration status.

### Statistical analysis: Before the pandemic

We compared pre-pandemic annual consultation rates between migrants and non-migrants using RRs derived from unadjusted and adjusted negative binomial models to account for overdispersion in the data. Covariates were individual’s time-varying age (i.e., their age in each study year represented by 5-year age categories), sex, study year and practice region, and the offset was the log person-years of follow-up. As socioeconomic status is commonly recognised as a mediator of the relationship between ethnicity and healthcare (23), we considered it a potentially important mediator of the effect of migration on consultation rates. We, therefore, ran multivariable models with and without adjusting for index of multiple deprivation (IMD; patient-level IMD for individuals with linked IMD data or else practice-level IMD). We stratified models by larger age groups (0–15, 16–24, 25–34, 35–49, 50–64, and 65 and over) to account for differences in representativeness of the migration code list across different age groups (20). Due to local policy interest, we conducted a secondary analysis of individuals in London only.

### Statistical analysis: Before versus during the pandemic

We explored any changes in the difference between migrants’ and non-migrants’ consultation rates before versus during the pandemic via an interrupted time series (ITS) analysis using a step-change model adapted from Mansfield et al. (14) (see Supplementary Box 2). We compared the pandemic period, defined as the time following introduction of national restrictions (29 March 2020 to 26 December 2020), with the pre-pandemic period (4 January 2015 to 7 March 2020). We added an interaction term between the pandemic period and migration status, which is interpreted as the additional effect that being a migrant versus a non-migrant had on consultation rates during the pandemic compared to any differences observed in the pre-pandemic period (i.e., the multiplicative effect of migration). We did not include a recovery slope term as our focus was on the average effect of migration on consultation rates during the initial months of the pandemic. We removed data for 8–28 March 2020 to account for behaviour changes in anticipation of pandemic restrictions. We matched migrants and non-migrants in the ITS cohort on a 1:1 ratio by sex, practice region, IMD and age at study start. In secondary analyses, we limited the analysis to London, and also stratified by consultation type (face-to-face and telephone) given considerable changes to service delivery during the pandemic.

### Statistical analysis: Effect modification by ethnicity

In the pre-pandemic analysis, we examined effect modification by ethnicity using a two-way interaction term between migration and ethnicity, based on the 2011 Census’ 18 categories grouped into 6 broader categories and derived using an ethnicity code list adapted from Pathak et al. (20), with an ‘Unknown’ category for individuals with no ethnicity data. We calculated the relative excess risk due to interaction (RERI, additive) (24) and multiplicative effects. We also calculated the effect of migration across each ethnicity strata, and compared migrants of each ethnicity to White British non-migrants (25). In the ITS analysis, we included a three-way interaction term between migration, ethnicity and pandemic to determine the effect of migration across each ethnicity strata (see Supplementary Box 2).

### Bias

We replaced the binary migrant status with the categorical migration certainty variable (‘definite’ and ‘probable’ migrants) to assess potential misclassification bias in the annual pre-pandemic and ITS analyses. To account for migrants’ younger age at cohort entry, later cohort entry, shorter time between entering the CPRD GOLD database and entering the cohort, and shorter follow-up time, we conducted two pre-pandemic and one ITS sensitivity analyses. First, we matched migrants and non-migrants on a 1:1 ratio by practice region, and age and year at entering CPRD GOLD. Second, we replaced the age and year at entering CPRD GOLD matching variables with follow-up time. Finally, we conducted an ITS sensitivity analysis by matching on age and year at entering CPRD GOLD (in addition to sex, practice region and IMD). We visually inspected RRs from the pre-pandemic annual analysis and ITS analysis for differences.

## Results

### Cohort characteristics

The pre-pandemic annual cohort comprised 601,033 individuals, of which 145,233 were migrants. Migrants were younger than non-migrants at cohort entry and exit, and their median time between entering CPRD GOLD and entering the study cohort was shorter (Table S3). These differences were generally attenuated in the matched annual-analysis cohorts (Tables S4 and S5).

The ITS cohort comprised 262,644 individuals, with half identified as migrants (Table 1). A greater proportion of migrants and non-migrants were located in London versus other regions. Fewer migrants were of White British ethnicity (3.2%) compared to non-migrants (44.6%). The proportion of White British non-migrants is lower than the national average (26), possibly due to unrecorded ethnicity for 31.7% of individuals. Migrants had a shorter time between CPRD GOLD entry and study entry compared with non-migrants (median of 0.4 and 3.8 years, respectively). However, matching migrants to non-migrants based on age and year of CPRD GOLD entry removed this difference (Table S6).

**Table 1:** Demographic characteristics of the ITS cohort (matched on age at study start, sex, practice region and IMD)

| Characteristic | Overall<br>N = 262,644 | Non-migrant<br>n = 131,322 (50.0%) | Migrant<br>n = 131,322 (50.0%) | Definite<br>n = 46,583 (35.5%) | Probable<br>n = 84,739 (64.5%) |
| --- | --- | --- | --- | --- | --- |
| *Follow up, person-years* |  |  |  |  |  |
| Total | 563,116 | 289,267 | 273,849 | 90,894 | 182,956 |
| Mean (SD) | 2.14 (1.89) | 2.20 (1.92) | 2.09 (1.86) | 1.95 (1.85) | 2.16 (1.85) |
| Median (IQR) | 1.55 (2.89) | 1.55 (2.95) | 1.55 (2.80) | 1.39 (2.78) | 1.65 (2.78) |
| *Sex, n (%)* |  |  |  |  |  |
| Male | 126,108 (48.0%) | 63,054 (48.0%) | 63,054 (48.0%) | 22,812 (49.0%) | 40,242 (47.5%) |
| Female | 136,536 (52.0%) | 68,268 (52.0%) | 68,268 (52.0%) | 23,771 (51.0%) | 44,497 (52.5%) |
| *Year of cohort entry, n (%)* |  |  |  |  |  |
| 2015 | 196,541 (74.8%) | 106,327 (81.0%) | 90,214 (68.7%) | 36,601 (78.6%) | 53,613 (63.3%) |
| 2016 | 17,470 (6.7%) | 6,642 (5.1%) | 10,828 (8.2%) | 3,326 (7.1%) | 7,502 (8.9%) |
| 2017 | 16,001 (6.1%) | 6,752 (5.1%) | 9,249 (7.0%) | 2,396 (5.1%) | 6,853 (8.1%) |
| 2018 | 14,520 (5.5%) | 5,256 (4.0%) | 9,264 (7.1%) | 2,112 (4.5%) | 7,152 (8.4%) |
| 2019 | 16,368 (6.2%) | 5,171 (3.9%) | 11,197 (8.5%) | 2,035 (4.4%) | 9,162 (10.8%) |
| 2020 | 1,744 (0.7%) | 1,174 (0.9%) | 570 (0.4%) | 113 (0.2%) | 457 (0.5%) |
| *Age at cohort entry, years* |  |  |  |  |  |
| Mean (SD) | 33 (18) | 33 (18) | 33 (18) | 35 (17) | 32 (19) |
| Median (IQR) | 32 (21) | 32 (21) | 32 (21) | 33 (20) | 32 (24) |
| *Age at cohort exit, years* |  |  |  |  |  |
| Mean (SD) | 35 (18) | 35 (18) | 35 (18) | 36 (17) | 34 (19) |
| Median (IQR) | 34 (22) | 34 (22) | 34 (22) | 35 (21) | 34 (24) |
| *Time between CPRD GOLD entry and study cohort entry, years* |  |  |  |  |  |
| Mean (SD) | 4.3 (5.8) | 6.2 (6.7) | 2.4 (3.8) | 2.5 (3.7) | 2.4 (3.8) |
| Median (IQR) | 1.8 (6.4) | 3.8 (11.3) | 0.4 (3.5) | 0.4 (3.7) | 0.4 (3.4) |
| *Ethnicity, n (%)* |  |  |  |  |  |
| White British | 62,762 (23.9%) | 58,561 (44.6%) | 4,201 (3.2%) | 1,883 (4.0%) | 2,318 (2.7%) |
| White non-British | 63,118 (24.0%) | 12,036 (9.2%) | 51,082 (38.9%) | 13,177 (28.3%) | 37,905 (44.7%) |
| Mixed/Multiple ethnic groups | 6,344 (2.4%) | 2,456 (1.9%) | 3,888 (3.0%) | 1,542 (3.3%) | 2,346 (2.8%) |
| Asian/Asian British | 43,126 (16.4%) | 7,415 (5.6%) | 35,711 (27.2%) | 9,858 (21.2%) | 25,853 (30.5%) |
| Black/African/Caribbean/Black British | 19,442 (7.4%) | 7,642 (5.8%) | 11,800 (9.0%) | 6,030 (12.9%) | 5,770 (6.8%) |
| Another ethnic group | 10,475 (4.0%) | 1,572 (1.2%) | 8,903 (6.8%) | 2,586 (5.6%) | 6,317 (7.5%) |
| Unknown | 57,377 (21.8%) | 41,640 (31.7%) | 15,737 (12.0%) | 11,507 (24.7%) | 4,230 (5.0%) |
| *Practice region, n (%)* |  |  |  |  |  |
| London | 100,020 (38.1%) | 50,010 (38.1%) | 50,010 (38.1%) | 22,003 (47.2%) | 28,007 (33.1%) |
| North East | 984 (0.4%) | 492 (0.4%) | 492 (0.4%) | 77 (0.2%) | 415 (0.5%) |
| North West | 28,982 (11.0%) | 14,491 (11.0%) | 14,491 (11.0%) | 4,769 (10.2%) | 9,722 (11.5%) |
| Yorkshire & The Humber | 612 (0.2%) | 306 (0.2%) | 306 (0.2%) | 73 (0.2%) | 233 (0.3%) |
| East Midlands | 64 (0.0%) | 32 (0.0%) | 32 (0.0%) | 10 (0.0%) | 22 (0.0%) |
| West Midlands | 20,080 (7.6%) | 10,040 (7.6%) | 10,040 (7.6%) | 1,333 (2.9%) | 8,707 (10.3%) |
| East of England | 14,326 (5.5%) | 7,163 (5.5%) | 7,163 (5.5%) | 957 (2.1%) | 6,206 (7.3%) |
| South West | 16,306 (6.2%) | 8,153 (6.2%) | 8,153 (6.2%) | 3,692 (7.9%) | 4,461 (5.3%) |
| South Central | 40,764 (15.5%) | 20,382 (15.5%) | 20,382 (15.5%) | 10,232 (22.0%) | 10,150 (12.0%) |
| South East Coast | 40,506 (15.4%) | 20,253 (15.4%) | 20,253 (15.4%) | 3,437 (7.4%) | 16,816 (19.8%) |
| *IMD, n (%)* |  |  |  |  |  |
| IMD 1 | 36,336 (13.8%) | 18,168 (13.8%) | 18,168 (13.8%) | 7,502 (16.1%) | 10,666 (12.6%) |
| IMD 2 | 38,976 (14.8%) | 19,488 (14.8%) | 19,488 (14.8%) | 6,116 (13.1%) | 13,372 (15.8%) |
| IMD 3 | 48,688 (18.5%) | 24,344 (18.5%) | 24,344 (18.5%) | 7,742 (16.6%) | 16,602 (19.6%) |
| IMD 4 | 66,990 (25.5%) | 33,495 (25.5%) | 33,495 (25.5%) | 11,548 (24.8%) | 21,947 (25.9%) |
| IMD 5 | 71,654 (27.3%) | 35,827 (27.3%) | 35,827 (27.3%) | 13,675 (29.4%) | 22,152 (26.1%) |
| *Patients in each study year, n (%)* |  |  |  |  |  |
| 2015 | 196,541 (27.5%) | 106,327 (29.4%) | 90,214 (25.6%) | 36,601 (31.2%) | 53,613 (22.8%) |
| 2016 | 138,899 (19.5%) | 73,028 (20.2%) | 65,871 (18.7%) | 24,212 (20.6%) | 41,659 (17.7%) |
| 2017 | 115,280 (16.1%) | 59,524 (16.5%) | 55,756 (15.8%) | 16,921 (14.4%) | 38,835 (16.5%) |
| 2018 | 103,295 (14.5%) | 49,249 (13.6%) | 54,046 (15.3%) | 16,314 (13.9%) | 37,732 (16.0%) |
| 2019 | 94,498 (13.2%) | 43,561 (12.1%) | 50,937 (14.5%) | 13,417 (11.4%) | 37,520 (16.0%) |
| 2020 | 65,296 (9.1%) | 29,697 (8.2%) | 35,599 (10.1%) | 9,857 (8.4%) | 25,742 (10.9%) |

### Before the pandemic

Migrants in the annual cohort attended fewer consultations than non-migrants; 4.31 (4.31– 4.32) compared with 5.62 (5.62–5.62) consultations per person-year (Table S7). A similar, although less pronounced, trend was observed in the pre-pandemic period of the ITS analysis (4.35 [4.34-4.36] versus 4.6 [4.59-4.6] consultations per person-year; Table S8).

Migrants had a 6% lower rate of consultations than non-migrants after multivariable adjustment (Figure 2A, RR:0.94, 95%CI:0.93-0.94). A similar RR was obtained when IMD was removed from the model (RR:0.95, 95%CI:0.94–0.95). Consultation rates in migrants were slightly higher than non-migrants for individuals aged 0–15 years, 50–64 years and 65 years and above, while migrants aged 16–24 years, 25–34 years and 35–49 years had lower consultation rates than non-migrants (Figure 2A).

**Figure 2:**
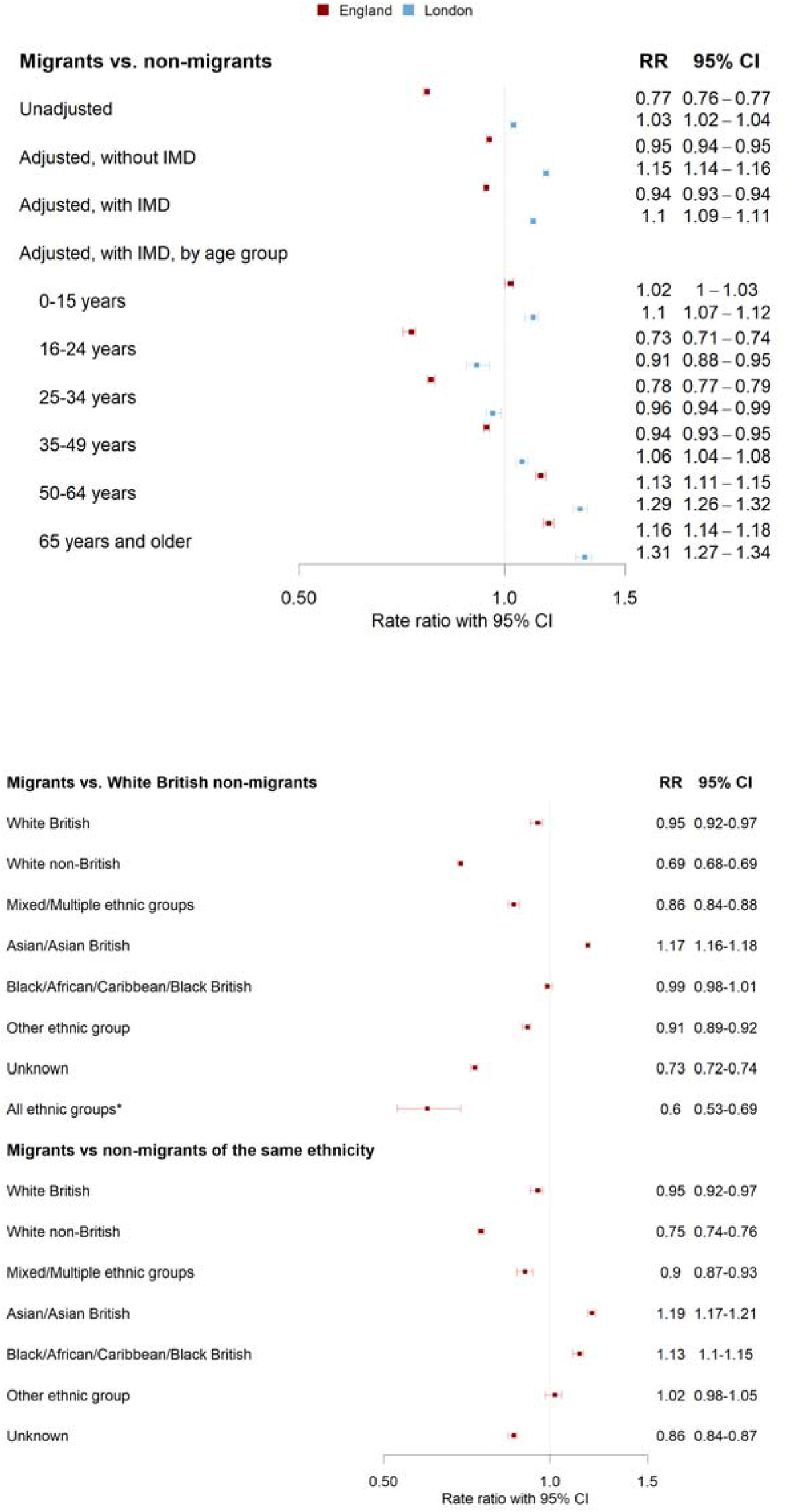
Forest plots of migrant versus non-migrant consultation rate ratios before the pandemic (2015-2019), including by age group (A) and ethnicity (B). *All represents migrants of all ethnic groups compared to White British non-migrants

In London, migrants had higher rates of consultation than non-migrants after multivariable adjustment (Figure 2A; RR:1.10, 95%CI:1.09-1.11). Consultation rates were only lower among migrants aged 16–24 years and 25–34 years; however, the differences in consultation rates between migrants and non-migrants in these age groups were not as great as those seen in the same age groups in England.

### Before versus during the pandemic

Crude face-to-face consultation rates reduced during the first eight months of the pandemic from 4.12 (4.11–4.13) to 3.02 (3–3.05) and 4.35 (4.34–4.36) to 3.52 (3.49–3.55) consultations per person-year for migrants and non-migrants, respectively (Table S8). Telephone consultations increased during the pandemic in both groups (from 0.23 [0.23– 0.23] to 0.52 [0.51–0.53] in migrants and 0.25 [0.25–0.25] to 0.68 [0.67–0.69] in non-migrants).

#### During the pandemic, migrants had lower rates of face-to-face (RR:0.86, 95%CI:0.81-0.9) and telephone consultations (RR:0.76, 95%CI:0.71-0.81) compared to non-migrants (Figure 3

Table 2). This resulted in an additional 9% reduction (RR:0.91, 95%CI:0.86–0.96) in the difference in face-to-face consultation rates observed between migrants and non-migrants pre-pandemic, and an additional 14% reduction for telephone consultations (RR:0.86, 95%CI:0.80–0.92).

**Table 2:**
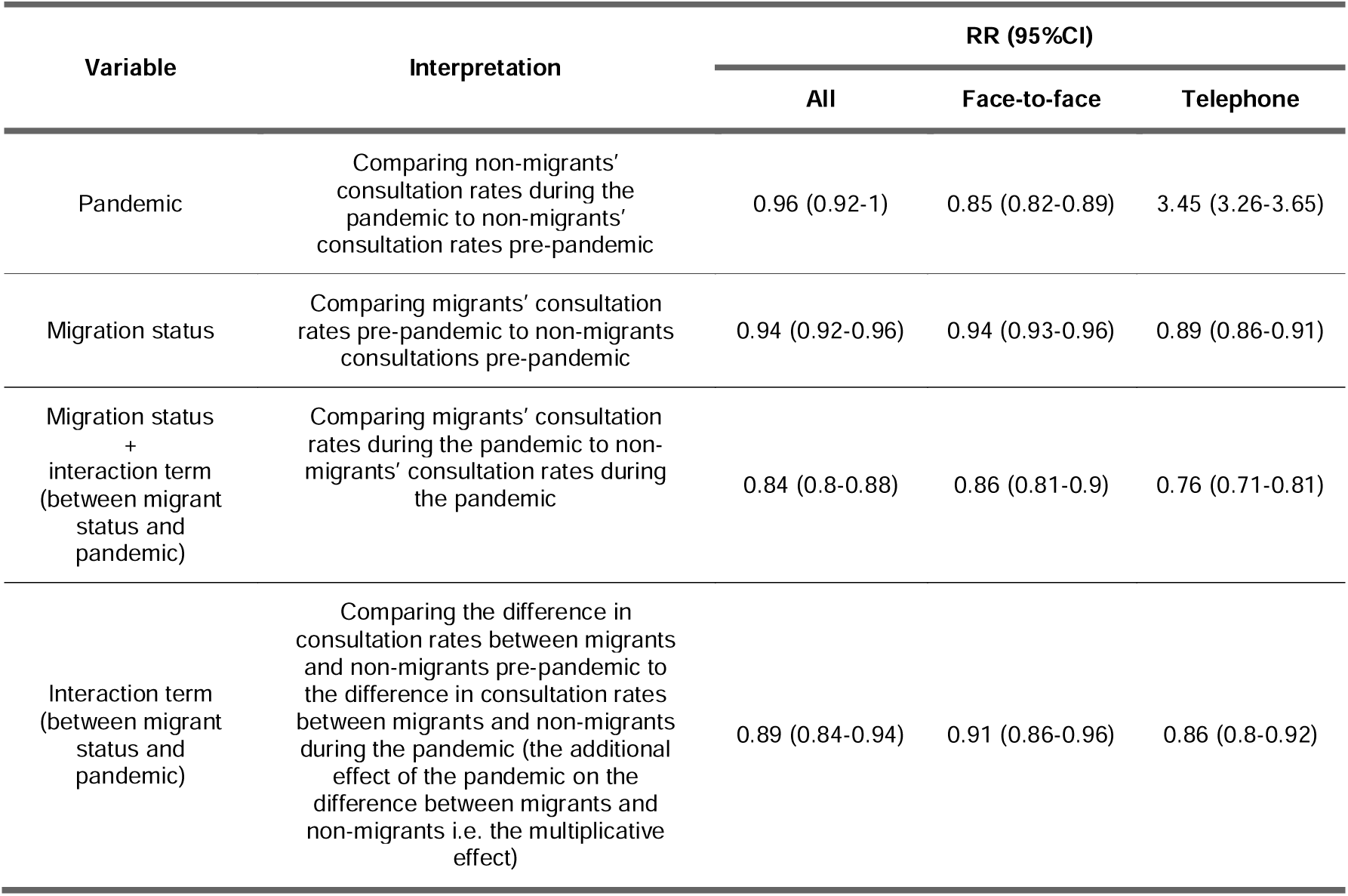
Consultation rate ratios from interrupted time-series analysis (5 January 2015 to 26 December 2020) for England

**Figure 3:**
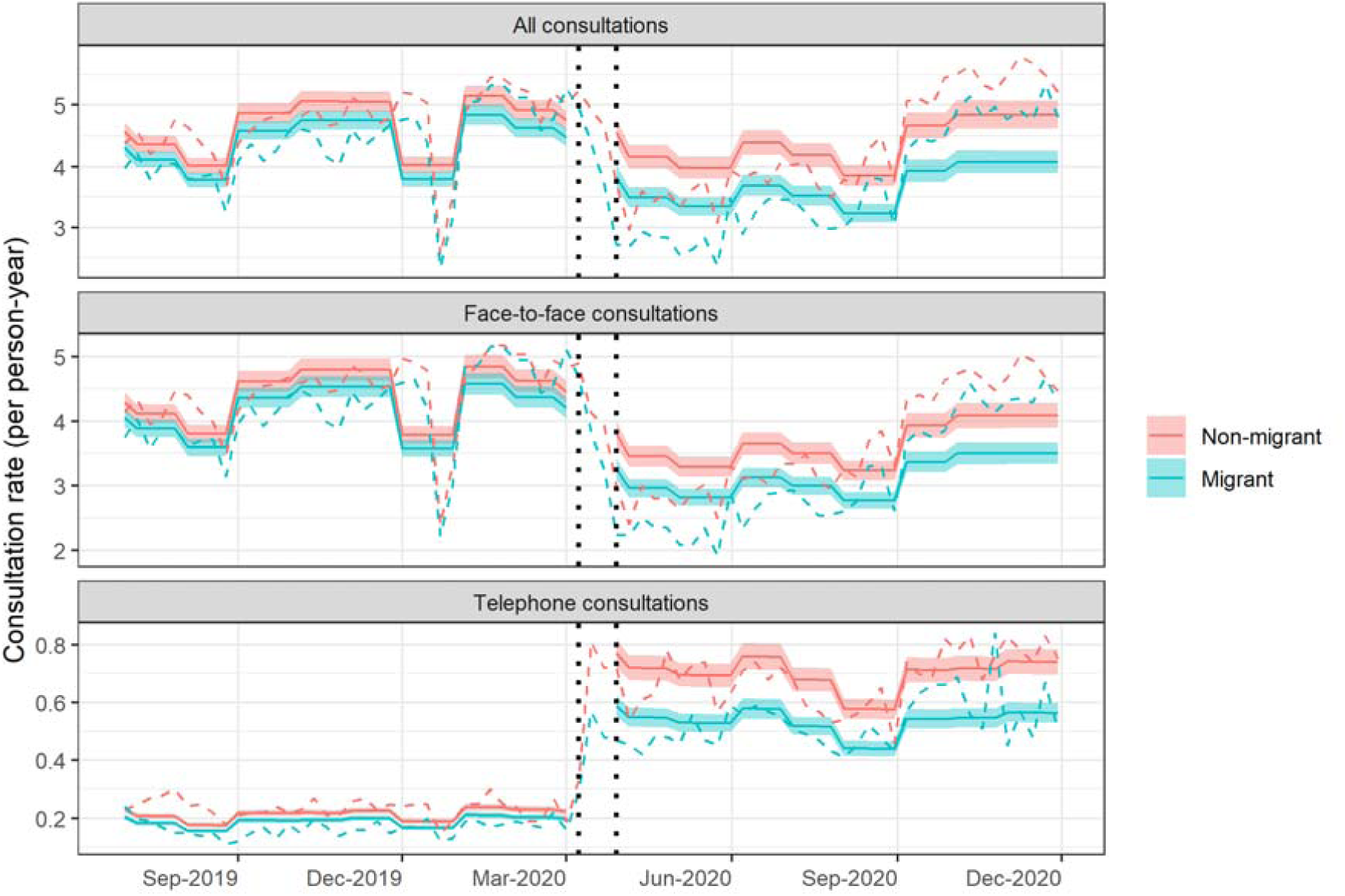
Weekly consultation rates by migration status in England: predicted rates from interrupted time-series analysis (solid line) and actual observed rates (dashed line), truncated view July 2019 - November 2020

This change was more pronounced in migrants aged 0–15 years, who attended more consultations before the pandemic (RR:1.05, 95%CI:1.02–1.07) and fewer during the pandemic (RR:0.76, 95%CI:0.71–0.82). This resulted in a 27% reduction in the pre-pandemic difference in consultation rates between migrants and non-migrants (RR:0.73, 95%CI:0.68–0.79). Other age groups were also negatively affected to varying degrees (Figure 4 and Table S9).

**Figure 4:**
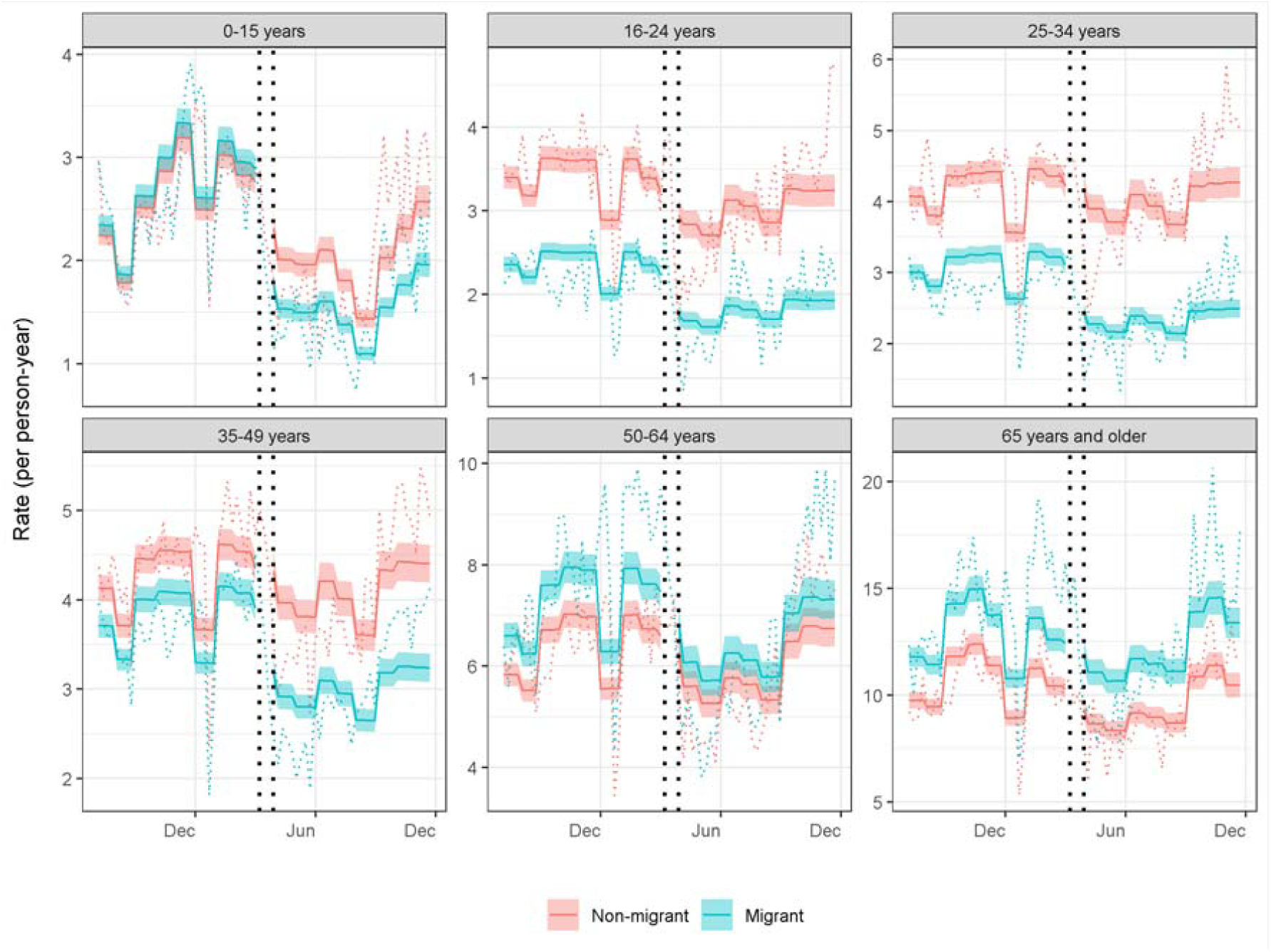
Weekly consultation rates by migration status and age group in England: predicted rates from interrupted time-series analysis (solid line) and actual observed rates (dashed line), truncated view July 2019– November 2020

In London, migrants had slightly higher rates both before and during the pandemic (Table S10). Migrants and non-migrants in London had similar rates of telephone consultations pre-pandemic (RR:0.99, 95%CI:0.95–1.03; Figure S1 and Table S10). However, during the pandemic migrants had substantially lower rates than non-migrants (RR:0.77, 95%CI:0.70– 0.86), which equates to a further 22% reduction compared with the pre-pandemic difference in consultation rates between migrants and non-migrants (RR:0.78, 95%CI:0.7–0.88).

### Effect modification by ethnicity

Pre-pandemic, White non-British migrants had the lowest consultation rates compared to non-migrants of the same ethnicity (Figure 2B; RR:0.75, 95%CI:0.74–0.76), followed by migrants of Unknown, Mixed/Multiple and White British ethnicities. Conversely, Asian/Asian British migrants (RR:1.19, 95%CI:1.17-1.21) and Black/African/Caribbean/Black British migrants (RR:1.13, 95%CI:1.1–1.15) had higher consultation rates than their non-migrant counterparts. Multiplicative and additive effects are presented in Table S11. For the majority of ethnicities, further disaggregation resulted in rate ratios that were consistent with the wider group estimate (Table S12). However, within the Mixed ethnic group, the lower consultation rate was primarily driven by individuals from Mixed White and Black Caribbean or African backgrounds, with no evidence of a difference between migrants and non-migrants of Mixed White and Asian or Other Mixed background.

Within ethnic groups, the largest additional reductions in consultation rates during the pandemic in migrants versus non-migrants were in the White British (RR:0.69, 95%CI:0.64– 0.73; Figure 5 and Table S13), Black/African/Caribbean/Black British (RR 0.68, 95%CI 0.64- 0.73), and White non-British (RR:0.72, 95%CI:0.68–0.77) backgrounds. Within the Black/African/Caribbean/Black British group the largest impact was in individuals of African ethnicity (Figure S2 and Table S14). In the Asian/Asian British ethnic group, the magnitude of the higher consultation rates observed pre-pandemic between migrants and non-migrants further increased during the pandemic (RR:1.11, 95%CI:1.04–1.18), which was driven by consultations in the Pakistani and Other Asian groups (Figure S2 and Table S14).

**Figure 5:**
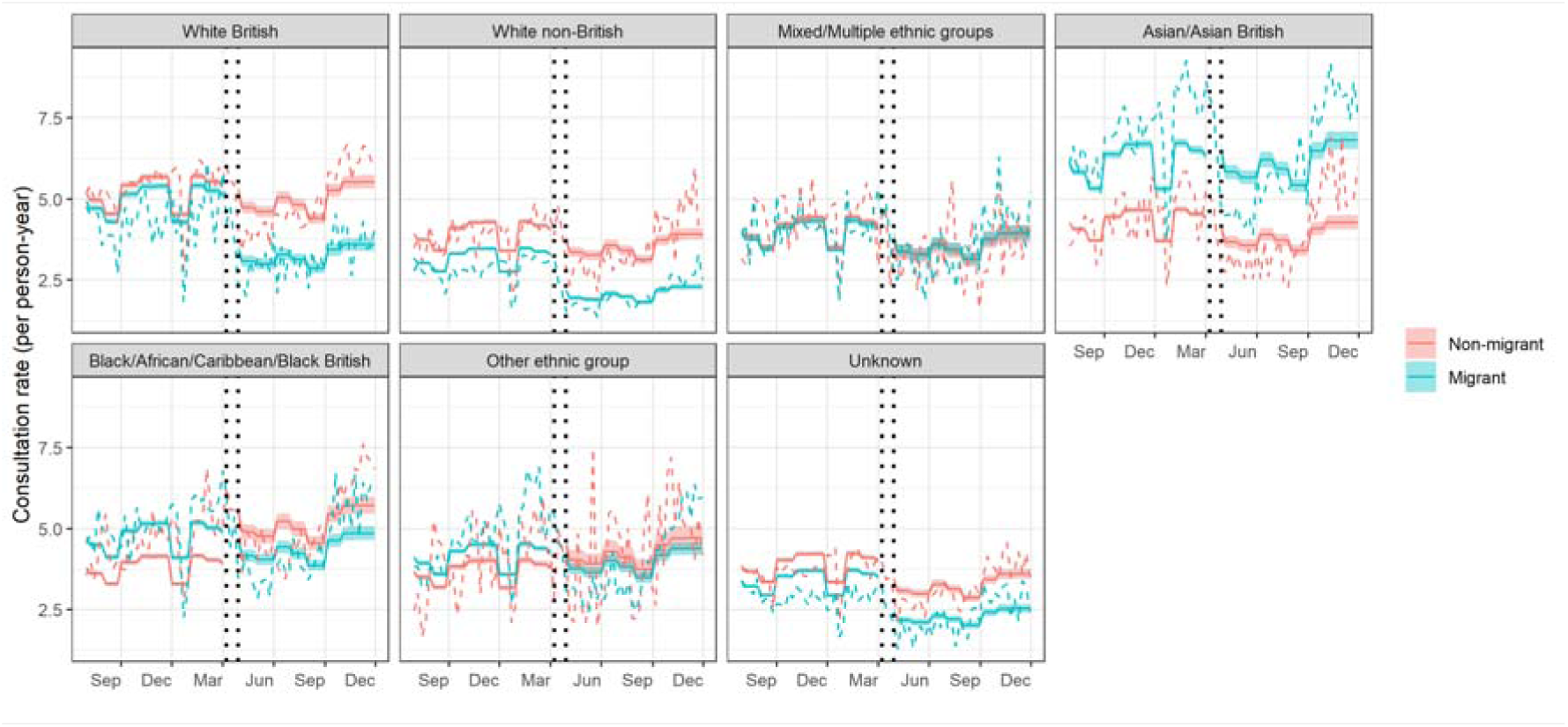
Weekly consultation rates by migration status and ethnicity in England: predicted rates from interrupted time-series analysis (solid line) and actual observed rates (dashed line), truncated view July 2019–November 2020

### Sensitivity analyses

In pre-pandemic sensitivity analyses (Figure S3), RRs were numerically lower than the main analysis RRs for both matched cohorts (i.e. [1] matched on age and year when an individual joined a CPRD practice and [2] matched on follow-up). When replacing binary migration status with migration certainty, ‘definite’ and ‘probable’ migrants only differed slightly (Figure S4).

Pre-pandemic estimates obtained from multivariable modelling largely corroborated findings from the ITS analysis over the same period, except in the sensitivity analysis where ‘definite’ migrants had higher rates. This is likely due to the lack of age adjustment in the ITS analysis from using aggregate data. The pandemic amplified the effect of migration on consultations for ‘probable’ migrants (RR:0.87, 95%CI:0.82–0.92) while the difference for ‘definite’ migrants (RR:0.96, 95%CI:0.91–1.02) was not significant (Figure S5 and Table S15).

Results from the ITS cohort sensitivity analysis were consistent with those of the main ITS analysis (Figure S6 and Table S16).

## Discussion

We present findings concerning migrants’ primary care utilisation in England using one of the most comprehensive sources of primary care research data available in the UK (21). We show that migrants had lower consultation rates than non-migrants before the COVID-19 pandemic and that the pandemic exacerbated this difference. Our findings also highlight that the effect of being a migrant on primary care utilisation varied significantly by age group and ethnicity.

Lower pre-pandemic consultation rates for migrants in this study are consistent with qualitative and survey-based studies of the multiple barriers that migrants face in accessing healthcare (3-7). Migrants in young and middle adulthood attended fewer consultations than non-migrants, while the opposite was seen in children and older adults. Higher rates in older adults aligns with previous research (12) and could be explained by the diminishing ‘healthy migrant effect’ over time (27) and increasing primary care consultation rates with time post-migration (11).

The exacerbation of differences in consultation rates between migrants and non-migrants during the pandemic was more prominent for telephone consultations than face-to-face consultations. It was also more pronounced for children, individuals of White British, White non-British and Black/African/Caribbean/Black British ethnicities, and individuals whose first language was not English. The large impact of the pandemic on migrant children and individuals from Black/African/Caribbean/Black British ethnic backgrounds could reflect known challenges accessing routine preventive care during the pandemic (28). Additionally, differences between telephone and face-to-face consultations and the impact on individuals whose first language was not English corroborates evidence from England that the shift from in-person to remote primary care exacerbated existing language and access barriers (18, 29). In the Asian/Asian British group, further increases during the pandemic to migrants’ already higher pre-pandemic consultation rates builds on existing pre-pandemic evidence of higher primary care use in individuals of South Asian ethnicity compared with other ethnic groups (30). Possible differences in healthcare needs underlying this finding within Asian/Asian British groups warrant further investigation.

In London, the large difference in telephone consultation rates between migrants and non-migrants that emerged during the pandemic could be due to changes in London’s migrant composition as a result of emigration (31). This composition change could have seen more migrants experiencing access barriers (e.g. digital and language barriers) remaining in London.

Limitations of our study include the under-recording of migration-related indicators in EHRs, which could result in migrants being misclassified as non-migrants (20) and a selection bias towards migrants who are more engaged with primary care. These biases would likely result in an underestimation of differences between groups. As we reported previously, the migration code list used in this study is less representative of migrants aged over 50 (20). As a result, findings concerning older migrants should be interpreted with caution. Another limitation is the lack of power in the 18-category ethnicity ITS analysis; these findings should also be interpreted with caution.

Our study could also have been affected by changes in size and composition of the migrant population during the study period due to the pandemic itself and/or other factors e.g. the UK’s exit from the EU. We found pronounced widening of differences between migrants’ and non-migrants’ consultation rates in both White British and White non-British groups, who may represent EU migrants. The lack of timely de-registration of migrants who leave GP practices and/or emigrate from the UK could contribute to a greater amount of false follow-up time (i.e. a numerator-denominator bias) and, thus, an underestimation of consultation rates. Finally, we provide quantitative evidence on migrants’ primary care utilisation, which is useful for service planning; however, we did not assess clinical need and, therefore, cannot make firm conclusions regarding the inequity of these differences.

As GP practices continue to use remote consultations (32), concerted efforts are needed to ensure all GP services are accessible to migrants. Clinical commissioning groups should address supply-side factors to support efficient and effective use of primary care (e.g. professional interpreting and translation services, culturally responsive service delivery plans) and demand-side factors (e.g. improving migrants’ knowledge of their healthcare entitlements and supporting migrants to make informed decisions about healthcare use during the pandemic and beyond) (18). Additionally, improvements are needed in both the completeness and accuracy of migration and ethnicity recording in primary care (33). Disaggregation of health outcomes by migrant sub-group is needed to better understand the needs of this diverse population (34) and inform service planning. However, socially excluded migrant sub-groups (e.g. asylum seekers, undocumented migrants, survivors of human trafficking) experience greater barriers accessing NHS services, fear data sharing for immigration enforcement purposes, and are thus rarely captured in routine health data (35). Further research is also needed to explore factors that affected migrants and non-migrants differently during the pandemic and whether differences during wave one have persisted or reduced to pre-pandemic levels in subsequent waves.

To conclude, the pandemic impacted migrants’ primary care usage more than that of non-migrants. Although our findings do not provide explanations for this disproportionate impact, they reinforce the need for health services to mitigate service-delivery related barriers and ensure migrants utilise primary care proportionate to their health needs. This requires policy-makers, commissioners and service planners to provide adequate resourcing for primary care to meet the diverse needs of their local migrant and ethnic communities across age groups. Further research is also needed to investigate whether changes in migrants’ primary care usage during the pandemic resulted in inequities in health outcomes.

## Notes

### Data sharing

This study used pseudonymised patient-level data from CPRD GOLD, which we are unable to publish to protect patient confidentiality. Other researchers can apply to use patient-level data in CPRD GOLD data through CPRD’s Research Data Governance Process (RDG; https://www.cprd.com/Data-access). All code used to generate these analyses is publicly available.

### Ethical approval

This study was approved by the UK Medicines and Healthcare products Regulatory Agency Independent Scientific Advisory Committee (protocol 19_062R, approval on 29 April 2019) and it was carried out as part of the CALIBER programme under Section 251 (NHS Social Care Act 2006), which has NHS research ethics approval (09/H0810/16).

## Data Availability

This study used pseudonymised patient-level data from CPRD GOLD, which we are unable to publish to protect patient confidentiality. Other researchers can apply to use patient-level data in CPRD GOLD data through CPRD's Research Data Governance Process (RDG; https://www.cprd.com/Data-access). All code used to generate these analyses is publicly available. All code for data cleaning and analysis is freely available.

https://doi.org/10.5281/zenodo.6345287

## Acknowledgements

This study is based in part on data from the Clinical Practice Research Datalink obtained under licence from the UK Medicines and Healthcare products Regulatory Agency. The data is provided by patients and collected by the NHS as part of their care and support. The interpretation and conclusions contained in this study are those of the authors alone. The use of ONS data is subject to copyright © (2021), re-used with the permission of The Health &Social Care Information Centre. All rights reserved.

This study was carried out as part of the CALIBER © resource (https://www.ucl.ac.uk/healthinformatics/caliber and https://www.caliberresearch.org/). CALIBER, led from the UCL Institute of Health Informatics, is a research resource providing validated electronic health record phenotyping algorithms and tools for national structured data sources.

We thank Dr Arturo Gonzales-Izquierdo, Muhammad Qummer Ul Arfeen, Natalie Fitzpatrick, Professor Spiros Denaxas, Nadia Elsay and Professor Henry Potts for data management, project and statistical support.

