## Supplementary Information for "Migrants’ primary care utilisation before and during the COVID-19 pandemic in England: An interrupted time series"

### ****Supplementary Box 1: Study cohort****

The up-to-standard (UTS) date indicates the date at which a practice is deemed to be contributing high quality data that are suitable for research use and is determined by two main factors^1^. Firstly, an assessment of the continuity of data recording by examining any meaningful gaps in the data provided by a practice. Secondly, an assurance that patients who have transferred out of the practice or who have died have not been removed from the data; this is assessed by examining gaps in death recording within a practice. The earliest date at which there are no meaningful gaps in the data and in death recording is the practices UTS date.

Patient exclusions were made if a patient’s follow-up start date began after their follow-up end date after applying the following exclusions; their certainty of migration using the migration phenotype was ‘possible’, i.e. determined by an ambiguous non-UK origin code such as “Asian origin”, which could refer to ethnicity or country of birth/origin; their age was 100 years and over, as numbers were very small and poorly representative of ONS population estimates^2^. Consultation events were excluded from the count if they were indirect administrative tasks, belonged to patients outside of the final included cohort or were recorded outside of an included patient’s follow-up period.

### ****Supplementary Box 2: Interrupted time–series statistical analysis****

The interrupted time-series analysis was carried out as reported by Mansfield et al^3^, with some adaptations. A negative binomial model was chosen due to the presence of overdispersion in the consultation count data, with a log offset by weekly total follow-up time for each group (migrants and non-migrants). The model comprised a binary pandemic term to determine the behaviour step–change, a binary migration status term to measure the effect of migration, and an interaction between migration status and the pandemic. This interaction term provides a comparison of the difference in consultation rates between migrants and non–migrants pre–pandemic to the difference in consultation rates between migrants and non–migrants during the pandemic. This is the equivalent of the additional effect of the pandemic on the difference between migrants and non–migrants i.e. the multiplicative effect of migration. Additional terms comprised a linear time variable to account for longer term trends in behaviour and a categorical calendar month variable to capture any seasonal effects. To account for autocorrelation, first–order lagged residuals were included in the model and the LJung–Box test^4^ was conducted to assess if there was evidence of autocorrelation following their inclusion. In cases where there was still evidence of autocorrelation after the addition of first–order lagged residuals, a partial autocorrelation function plot was inspected, and the appropriate order of lagged residuals were added to the model.

To assess the effect of migration and ethnicity on consultation rates pre- and during the pandemic, the following additional interaction terms were included in the model: (1) an interaction between ethnicity and the pandemic and (2) a three-way interaction between migrant status, ethnicity and the pandemic. Using this model, the effect of being a migrant versus a non–migrant from the same ethnic group on the consultation rate during compared to before the pandemic could ascertained (represented by combining the migrant status/pandemic interaction term with the three–way migrant status/ethnicity/pandemic interaction term).

### Table S1· Categorisation of CPRD consultation types

| **Direct consultations** | | **Indirect administrative activities** |
| --- | --- | --- |
| ***Face–to–face consultation*** | ***Phone consultation*** |  |
| 1 Clinic | 21 Telephone call to a patient | 0 Data Not Entered |
| 2 Night visit, Deputising service | 35 Co–op Telephone advice | 5 Mail from patient |
| 3 Follow–up/routine visit | 55 Telephone Consultation | 10 Telephone call from a patient |
| 4 Night visit, Local rota |  | 12 Discharge details |
| 6 Night visit, practice |  | 13 Letter from Outpatients |
| 7 Out of hours, Practice |  | 14 Repeat Issue |
| 8 Out of hours, Non Practice |  | 15 Other |
| 9 Surgery consultation |  | 16 Results recording |
| 11 Acute visit |  | 17 Mail to patient |
| 18 Emergency Consultation |  | 19 Administration |
| 20 Casualty Attendance |  | 22 Third Party Consultation |
| 24 Children's Home Visit |  | 23 Hospital Admission |
| 27 Home Visit |  | 25 Day Case Report |
| 28 Hotel Visit |  | 26 GOS18 Report |
| 30 Nursing Home Visit |  | 29 NHS Direct Report |
| 31 Residential Home Visit |  | 33 Triage |
| 32 Twilight Visit |  | 39 Medicine Management |
| 34 Walk–in Centre |  | 41 Community Nursing Note |
| 36 Co–op Surgery Consultation |  | 42 Community Nursing Report |
| 37 Co–op Home Visit |  | 43 Data Transferred from other system |
| 38 Minor Injury Service |  | 44 Health Authority Entry |
| 40 Community Clinic |  | 45 Health Visitor Note |
| 48 Initial Post Discharge Review |  | 46 Health Visitor Report |
| 50 Night Visit |  | 47 Hospital Inpatient Report |
| 61 Extended Hours |  | 49 Laboratory Request |
|  |  | 51 Radiology Request |
|  |  | 52 Radiology Result |
|  |  | 53 Referral Letter |
|  |  | 54 Social Services Report |
|  |  | 56 Template Entry |
|  |  | 57 GP to GP communication transaction |
|  |  | 58 Non–consultation medication data |
|  |  | 59 Non–consultation data |
|  |  | 60 ePharmacy message |

Adapted from [https://caliberresearch·org/portal/show/clinical_contact](https://caliberresearch.org/portal/show/clinical_contact) and [https://caliberresearch·org/portal/show/consult](https://caliberresearch.org/portal/show/consult)

### Table S2· Ethnicity code list

| **6 level grouping** | **18 level grouping** | **Read code** | **Read term** | **Medcode** |
| --- | --- | --- | --- | --- |
| White British | British | 9S10·00 | White British | 12446 |
| White British | British | 9S13·00 | White Scottish | 26467 |
| White British | British | 9S14·00 | Other white British ethnic group | 26310 |
| White British | British | 9i00·00 | White British – ethnic category 2001 census | 98111 |
| White British | British | 9i20·00 | English – ethnic category 2001 census | 12352 |
| White British | British | 9i21·00 | Scottish – ethnic category 2001 census | 12436 |
| White British | British | 9i22·00 | Welsh – ethnic category 2001 census | 12681 |
| White British | British | 9i23·00 | Cornish – ethnic category 2001 census | 28887 |
| White British | British | 9t20·00 | White: Scottish – Scotland ethnic category 2011 census | 110432 |
| White British | British | 9t21·00 | White: other British – Scotland ethnic category 2011 census | 110694 |
| White British | British | 9t00·00 | White:Eng/Welsh/Scot/NI/Brit – England and Wales 2011 census | 110420 |
| White British | British | 9i24·00 | Northern Irish – ethnic category 2001 census | 42294 |
| White British | British | 9t10·00 | White – Northern Ireland ethnic category 2011 census | 112899 |
| White British | British | 9i0··00 | British or mixed British – ethnic category 2001 census | 12351 |
| White Non–British | Irish | 9S11·00 | White Irish | 24837 |
| White Non–British | Irish | 9SA9·00 | Irish (NMO) | 24270 |
| White Non–British | Irish | 9i1··00 | Irish – ethnic category 2001 census | 12532 |
| White Non–British | Irish | 9i10·00 | White Irish – ethnic category 2001 census | 98213 |
| White Non–British | Irish | 9t22·00 | White: Irish – Scotland ethnic category 2011 census | 110687 |
| White Non–British | Irish | 9t01·00 | White: Irish – England and Wales ethnic category 2011 census | 110556 |
| White Non–British | Irish | 9i2Q·00 | Mixed Irish and other White – ethnic category 2001 census | 26391 |
| White Non–British | Gypsy or Irish Traveller | 9SI··00 | Irish traveller | 47601 |
| White Non–British | Gypsy or Irish Traveller | 9i2C·00 | Irish Traveller – ethnic category 2001 census | 55223 |
| White Non–British | Gypsy or Irish Traveller | 9i2D·00 | Traveller – ethnic category 2001 census | 55113 |
| White Non–British | Gypsy or Irish Traveller | 9i2E·00 | Gypsy/Romany – ethnic category 2001 census | 42290 |
| White Non–British | Gypsy or Irish Traveller | 9t02·00 | White: Gypsy/Irish Traveller – Eng+Wales eth cat 2011 census | 111386 |
| White Non–British | Gypsy or Irish Traveller | 9t23·00 | White: Gypsy/Irish Traveller – Scotland ethnic cat 2011 cens | 113253 |
| White Non–British | Gypsy or Irish Traveller | 9T2··00 | Traveller – gypsy | 32781 |
| White Non–British | Other White | 9S12·00 | Other white ethnic group | 12444 |
| White Non–British | Other White | 9SAA·00 | Greek/Greek Cypriot (NMO) | 45947 |
| White Non–British | Other White | 9SAA·11 | Greek (NMO) | 45955 |
| White Non–British | Other White | 9SAA·12 | Greek Cypriot (NMO) | 47949 |
| White Non–British | Other White | 9SAB·00 | Turkish/Turkish Cypriot (NMO) | 32066 |
| White Non–British | Other White | 9SAB·11 | Turkish (NMO) | 32126 |
| White Non–British | Other White | 9SAB·12 | Turkish Cypriot (NMO) | 32069 |
| White Non–British | Other White | 9SAC·00 | Other European (NMO) | 12633 |
| White Non–British | Other White | 9i2··00 | Other White background – ethnic category 2001 census | 12421 |
| White Non–British | Other White | 9i26·00 | Cypriot (part not stated) – ethnic category 2001 census | 32778 |
| White Non–British | Other White | 9i27·00 | Greek – ethnic category 2001 census | 12355 |
| White Non–British | Other White | 9i28·00 | Greek Cypriot – ethnic category 2001 census | 12769 |
| White Non–British | Other White | 9i29·00 | Turkish – ethnic category 2001 census | 12746 |
| White Non–British | Other White | 9i2A·00 | Turkish Cypriot – ethnic category 2001 census | 32413 |
| White Non–British | Other White | 9i2B·00 | Italian – ethnic category 2001 census | 12412 |
| White Non–British | Other White | 9i2F·00 | Polish – ethnic category 2001 census | 12467 |
| White Non–British | Other White | 9i2G·00 | Baltic Estonian/Latvian/Lithuanian – ethn categ 2001 census | 12433 |
| White Non–British | Other White | 9i2H·00 | Commonwealth (Russian) Indep States – ethn categ 2001 census | 28973 |
| White Non–British | Other White | 9i2J·00 | Kosovan – ethnic category 2001 census | 26341 |
| White Non–British | Other White | 9i2K·00 | Albanian – ethnic category 2001 census | 25422 |
| White Non–British | Other White | 9i2L·00 | Bosnian – ethnic category 2001 census | 46956 |
| White Non–British | Other White | 9i2M·00 | Croatian – ethnic category 2001 census | 28866 |
| White Non–British | Other White | 9i2N·00 | Serbian – ethnic category 2001 census | 47074 |
| White Non–British | Other White | 9i2P·00 | Other republics former Yugoslavia – ethnic categ 2001 census | 28936 |
| White Non–British | Other White | 9i2T·00 | Other White or White unspecified ethnic category 2001 census | 12591 |
| White Non–British | Other White | 9S1··00 | White | 22467 |
| White Non–British | Other White | 9i25·00 | Ulster Scots – ethnic category 2001 census | 40102 |
| White Non–British | Other White | 9i2R·00 | Oth White European/European unsp/Mixed European 2001 census | 12402 |
| White Non–British | Other White | 9i2S·00 | Other mixed White – ethnic category 2001 census | 28900 |
| White Non–British | Other White | 9t24·00 | White: Polish – Scotland ethnic category 2011 census | 110465 |
| White Non–British | Other White | 9t03·00 | White: other White backgrd– Eng+Wales ethnic cat 2011 census | 110407 |
| White Non–British | Other White | 9t25·00 | White: other White ethnic grp– Scotland ethnic cat 2011 cens | 110695 |
| White Non–British | Other White | 9T11·00 | New Zealand European | 57286 |
| White Non–British | Other White | 9T12·00 | Other European in New Zealand | 85505 |
| White Non–British | Other White | 9T7··00 | Slovak | 101787 |
| White Non–British | Other White | 134B·00 | RACE: Caucasian | 12191 |
| White Non–British | Other White | 134N·00 | RACE: White | 25550 |
| Mixed/Multiple ethnic group | Mixed White and Black Caribbean | 9SB5·00 | Black Caribbean and White | 32425 |
| Mixed/Multiple ethnic group | Mixed White and Black Caribbean | 9i3··00 | White and Black Caribbean – ethnic category 2001 census | 12742 |
| Mixed/Multiple ethnic group | Mixed White and Black Caribbean | 9t12·00 | Mixed: White and Black Caribbean – NI ethnic cat 2011 census | 110661 |
| Mixed/Multiple ethnic group | Mixed White and Black Caribbean | 9t04·00 | Mixed: White+Black Caribbean – Eng+Wales eth cat 2011 census | 110445 |
| Mixed/Multiple ethnic group | Mixed White and Black African | 9SB6·00 | Black African and White | 32443 |
| Mixed/Multiple ethnic group | Mixed White and Black African | 9i4··00 | White and Black African – ethnic category 2001 census | 12437 |
| Mixed/Multiple ethnic group | Mixed White and Black African | 9t13·00 | Mixed: White and Black African – NI ethnic cat 2011 census | 110651 |
| Mixed/Multiple ethnic group | Mixed White and Black African | 9t05·00 | Mixed: White+Black African – Eng+Wales eth cat 2011 census | 110421 |
| Mixed/Multiple ethnic group | Mixed White and Black African | 134L·00 | RACE: Afro–caucasian | 45167 |
| Mixed/Multiple ethnic group | Mixed White and Asian | 9SB2·00 | Other ethnic, Asian/White orig | 32401 |
| Mixed/Multiple ethnic group | Mixed White and Asian | 9i5··00 | White and Asian – ethnic category 2001 census | 12638 |
| Mixed/Multiple ethnic group | Mixed White and Asian | 9i63·00 | Chinese and White – ethnic category 2001 census | 12706 |
| Mixed/Multiple ethnic group | Mixed White and Asian | 9t14·00 | Mixed: White and Asian – NI ethnic category 2011 census | 110471 |
| Mixed/Multiple ethnic group | Mixed White and Asian | 9t06·00 | Mixed: White+Asian – Eng+Wales ethnic category 2011 census | 110652 |
| Mixed/Multiple ethnic group | Other Mixed | 9SB3·00 | Other ethnic, mixed white orig | 35459 |
| Mixed/Multiple ethnic group | Other Mixed | 9i60·00 | Black and Asian – ethnic category 2001 census | 12795 |
| Mixed/Multiple ethnic group | Other Mixed | 9i61·00 | Black and Chinese – ethnic category 2001 census | 49940 |
| Mixed/Multiple ethnic group | Other Mixed | 9SB··00 | Other ethnic, mixed origin | 12696 |
| Mixed/Multiple ethnic group | Other Mixed | 9SB4·00 | Other ethnic, other mixed orig | 32420 |
| Mixed/Multiple ethnic group | Other Mixed | 9i6··00 | Other Mixed background – ethnic category 2001 census | 12873 |
| Mixed/Multiple ethnic group | Other Mixed | 9i64·00 | Asian and Chinese – ethnic category 2001 census | 47005 |
| Mixed/Multiple ethnic group | Other Mixed | 9i65·00 | Other Mixed or Mixed unspecified ethnic category 2001 census | 32408 |
| Mixed/Multiple ethnic group | Other Mixed | 9S51·00 | Other Black – Black/White orig | 25623 |
| Mixed/Multiple ethnic group | Other Mixed | 9SB1·00 | Other ethnic, Black/White orig | 47401 |
| Mixed/Multiple ethnic group | Other Mixed | 9i62·00 | Black and White – ethnic category 2001 census | 40110 |
| Mixed/Multiple ethnic group | Other Mixed | 9iA9·00 | Mixed Asian – ethnic category 2001 census | 46056 |
| Mixed/Multiple ethnic group | Other Mixed | 9iD3·00 | Mixed Black – ethnic category 2001 census | 40096 |
| Mixed/Multiple ethnic group | Other Mixed | 9t15·00 | Mixed: other Mixed/multiple ethnic backgrd – NI 2011 census | 110536 |
| Mixed/Multiple ethnic group | Other Mixed | 9t26·00 | Mixed/multiple ethnic grps: any– Scot ethnic cat 2011 census | 110696 |
| Mixed/Multiple ethnic group | Other Mixed | 9t07·00 | Mixed: other Mixed/multiple backgrd – Eng+Wales 2011 census | 110654 |
| Mixed/Multiple ethnic group | Other Mixed | 9S52·00 | Other Black – Black/Asian orig | 32165 |
| Mixed/Multiple ethnic group | Other Mixed | 9S5··00 | Black – other, mixed | 25676 |
| Mixed/Multiple ethnic group | Other Mixed | 134J·00 | RACE: Mixed | 30408 |
| Asian/Asian British | Indian | 9S6··00 | Indian | 12482 |
| Asian/Asian British | Indian | 9SA7·00 | Indian sub–continent (NMO) | 39696 |
| Asian/Asian British | Indian | 9i7··00 | Indian or British Indian – ethnic category 2001 census | 12414 |
| Asian/Asian British | Indian | 9t16·00 | Asian or Asian British: Indian – NI ethnic cat 2011 census | 110422 |
| Asian/Asian British | Indian | 9t28·00 | Asian: Indian, Indian Scot/Indian Brit– Scotland 2011 census | 111368 |
| Asian/Asian British | Indian | 9t08·00 | Asian/Asian Brit: Indian – Eng+Wales ethnic cat 2011 census | 110477 |
| Asian/Asian British | Indian | 9T1D·00 | Indian | 25920 |
| Asian/Asian British | Pakistani | 9S7··00 | Pakistani | 24690 |
| Asian/Asian British | Pakistani | 9i8··00 | Pakistani or British Pakistani – ethnic category 2001 census | 12460 |
| Asian/Asian British | Pakistani | 9t09·00 | Asian/Asian British:Pakistani– Eng+Wales eth cat 2011 census | 110464 |
| Asian/Asian British | Pakistani | 9t17·00 | Asian/Asian British: Pakistani – NI ethnic cat 2011 census | 110538 |
| Asian/Asian British | Pakistani | 9t27·00 | Asian: Pakistani/Pakistani Scot/Pakistani Brit– Scot 2011 | 110460 |
| Asian/Asian British | Pakistani | 134M·00 | RACE: Pakistani | 26062 |
| Asian/Asian British | Bangladeshi | 9S8··00 | Bangladeshi | 24740 |
| Asian/Asian British | Bangladeshi | 9i9··00 | Bangladeshi or British Bangladeshi – ethn categ 2001 census | 28888 |
| Asian/Asian British | Bangladeshi | 9t18·00 | Asian/Asian British: Bangladeshi – NI ethnic cat 2011 census | 110720 |
| Asian/Asian British | Bangladeshi | 9t0A·00 | Asian/Asian Brit: Bangladeshi– Eng+Wales eth cat 2011 census | 110590 |
| Asian/Asian British | Bangladeshi | 9t29·00 | Bangladeshi, Bangladeshi Scot or Bangladeshi Brit– Scot 2011 | 112225 |
| Asian/Asian British | Bangladeshi | 134I·00 | RACE: Bangladeshi | 26348 |
| Asian/Asian British | Chinese | 9S9··00 | Chinese | 24272 |
| Asian/Asian British | Chinese | 9iE··00 | Chinese – ethnic category 2001 census | 12468 |
| Asian/Asian British | Chinese | 9t2A·00 | Asian: Chinese – Scotland ethnic category 2011 census | 111064 |
| Asian/Asian British | Chinese | 9t19·00 | Asian/Asian British: Chinese – NI ethnic cat 2011 census | 112363 |
| Asian/Asian British | Chinese | 9t0B·00 | Asian/Asian Brit: Chinese – Eng+Wales ethnic cat 2011 census | 110922 |
| Asian/Asian British | Chinese | 9T1C·00 | Chinese | 12718 |
| Asian/Asian British | Chinese | 134D·00 | RACE: Chinese | 32224 |
| Asian/Asian British | Other Asian | 9iA7·00 | Caribbean Asian – ethnic category 2001 census | 32399 |
| Asian/Asian British | Other Asian | 9iA1·00 | Punjabi – ethnic category 2001 census | 26392 |
| Asian/Asian British | Other Asian | 9SA6·00 | E Afric Asian/Indo–Carib (NMO) | 38097 |
| Asian/Asian British | Other Asian | 9iA2·00 | Kashmiri – ethnic category 2001 census | 64133 |
| Asian/Asian British | Other Asian | 9SA6·11 | East African Asian (NMO) | 46818 |
| Asian/Asian British | Other Asian | 9SA8·00 | Other Asian (NMO) | 26379 |
| Asian/Asian British | Other Asian | 9SH··00 | Other Asian ethnic group | 12668 |
| Asian/Asian British | Other Asian | 9iA··00 | Other Asian background – ethnic category 2001 census | 12513 |
| Asian/Asian British | Other Asian | 9iA3·00 | East African Asian – ethnic category 2001 census | 47077 |
| Asian/Asian British | Other Asian | 9iA8·00 | British Asian – ethnic category 2001 census | 12653 |
| Asian/Asian British | Other Asian | 9iAA·00 | Other Asian or Asian unspecified ethnic category 2001 census | 28935 |
| Asian/Asian British | Other Asian | 9SA6·12 | Indo–Caribbean (NMO) | 99316 |
| Asian/Asian British | Other Asian | 9t2B·00 | Asian: other Asian group – Scotland ethnic cat 2011 census | 110855 |
| Asian/Asian British | Other Asian | 9t0C·00 | Asian/Asian Brit: other Asian– Eng+Wales eth cat 2011 census | 111743 |
| Asian/Asian British | Other Asian | 9t1A·00 | Asian/Asian British: other Asian – NI ethnic cat 2011 census | 110425 |
| Asian/Asian British | Other Asian | 9T1E·00 | Other Asian | 32396 |
| Asian/Asian British | Other Asian | 9iA4·00 | Sri Lankan – ethnic category 2001 census | 12608 |
| Asian/Asian British | Other Asian | 9iA5·00 | Tamil – ethnic category 2001 census | 12760 |
| Asian/Asian British | Other Asian | 9iA6·00 | Sinhalese – ethnic category 2001 census | 12887 |
| Asian/Asian British | Other Asian | 9iF0·00 | Vietnamese – ethnic category 2001 census | 12719 |
| Asian/Asian British | Other Asian | 9iF1·00 | Japanese – ethnic category 2001 census | 12473 |
| Asian/Asian British | Other Asian | 9iF2·00 | Filipino – ethnic category 2001 census | 12420 |
| Asian/Asian British | Other Asian | 9iF3·00 | Malaysian – ethnic category 2001 census | 12730 |
| Asian/Asian British | Other Asian | 9T1B·00 | South East Asian | 46649 |
| Asian/Asian British | Other Asian | 9SC··00 | Vietnamese | 25411 |
| Asian/Asian British | Other Asian | 9T9··00 | Nepali | 101162 |
| Asian/Asian British | Other Asian | 134E·00 | RACE: Japanese | 45163 |
| Asian/Asian British | Other Asian | 134G·00 | RACE: Oriental | 41200 |
| Asian/Asian British | Other Asian | 134F·00 | RACE: Korean | 41394 |
| Black/African/Caribbean/Black British | African | 9S3··00 | Black African | 12778 |
| Black/African/Caribbean/Black British | African | 9S44·00 | Black – other African country | 35412 |
| Black/African/Caribbean/Black British | African | 9SA5·00 | Other African countries (NMO) | 47969 |
| Black/African/Caribbean/Black British | African | 9iC··00 | African – ethnic category 2001 census | 12350 |
| Black/African/Caribbean/Black British | African | 9iD1·00 | Nigerian – ethnic category 2001 census | 32886 |
| Black/African/Caribbean/Black British | African | 9S43·11 | Black North African | 46812 |
| Black/African/Caribbean/Black British | African | 9t2D·00 | African: any other African – Scotland ethnic cat 2011 census | 110655 |
| Black/African/Caribbean/Black British | African | 9t1B·00 | Black/Afri/Carib/Black Brit: African– NI eth cat 2011 census | 110630 |
| Black/African/Caribbean/Black British | African | 9t2C·00 | African: African/African Scot/African Brit – Scotland 2011 | 111059 |
| Black/African/Caribbean/Black British | African | 9t0D·00 | Black/African/Carib/Black Brit: African– Eng+Wales 2011 cens | 110437 |
| Black/African/Caribbean/Black British | African | 9iFA·00 | North African – ethnic category 2001 census | 47028 |
| Black/African/Caribbean/Black British | Caribbean | 9S2··00 | Black Caribbean | 12632 |
| Black/African/Caribbean/Black British | Caribbean | 9S42·00 | Black Caribbean/W·I·/Guyana | 57435 |
| Black/African/Caribbean/Black British | Caribbean | 9S42·11 | Black Caribbean | 47950 |
| Black/African/Caribbean/Black British | Caribbean | 9S42·12 | Black West Indian | 47997 |
| Black/African/Caribbean/Black British | Caribbean | 9S42·13 | Black Guyana | 32100 |
| Black/African/Caribbean/Black British | Caribbean | 9SA3·00 | Caribbean I·/W·I·/Guyana (NMO) | 54593 |
| Black/African/Caribbean/Black British | Caribbean | 9SA3·11 | Caribbean Island (NMO) | 57094 |
| Black/African/Caribbean/Black British | Caribbean | 9SA3·12 | West Indian (NMO) | 57075 |
| Black/African/Caribbean/Black British | Caribbean | 9SA3·13 | Guyana (NMO) | 93144 |
| Black/African/Caribbean/Black British | Caribbean | 9iB··00 | Caribbean – ethnic category 2001 census | 12432 |
| Black/African/Caribbean/Black British | Caribbean | 9t0E·00 | Black/African/Caribbn/Black Brit: Caribbean – Eng+Wales 2011 | 110436 |
| Black/African/Caribbean/Black British | Caribbean | 9t1C·00 | Black/Afri/Carib/Black Brit: Caribbean– NI eth cat 2011 cens | 110779 |
| Black/African/Caribbean/Black British | Caribbean | 9t2E·00 | Carib/Black: Caribbean/Carib Scot/Carib Brit– Scotland 2011 | 113671 |
| Black/African/Caribbean/Black British | Caribbean | 134K·00 | RACE: West indian | 32095 |
| Black/African/Caribbean/Black British | Caribbean | 134H·00 | RACE: Afro–caribbean | 25894 |
| Black/African/Caribbean/Black British | Other Black | 9S41·00 | Black British | 12452 |
| Black/African/Caribbean/Black British | Other Black | 9iD2·00 | Black British – ethnic category 2001 census | 40097 |
| Black/African/Caribbean/Black British | Other Black | 9S45·00 | Black E Afric Asia/Indo–Caribb | 47965 |
| Black/African/Caribbean/Black British | Other Black | 9S45·11 | Black East African Asian | 57753 |
| Black/African/Caribbean/Black British | Other Black | 9S45·12 | Black Indo–Caribbean | 57763 |
| Black/African/Caribbean/Black British | Other Black | 9S46·00 | Black Indian sub–continent | 48005 |
| Black/African/Caribbean/Black British | Other Black | 9S47·00 | Black – other Asian | 35350 |
| Black/African/Caribbean/Black British | Other Black | 9iD0·00 | Somali – ethnic category 2001 census | 12443 |
| Black/African/Caribbean/Black British | Other Black | 9S4··00 | Black, other, non–mixed origin | 24339 |
| Black/African/Caribbean/Black British | Other Black | 9S43·13 | Black Iranian | 50286 |
| Black/African/Caribbean/Black British | Other Black | 9S48·00 | Black – other | 26312 |
| Black/African/Caribbean/Black British | Other Black | 9SG··00 | Other black ethnic group | 32136 |
| Black/African/Caribbean/Black British | Other Black | 9iD··00 | Other Black background – ethnic category 2001 census | 32389 |
| Black/African/Caribbean/Black British | Other Black | 9iD4·00 | Other Black or Black unspecified ethnic category 2001 census | 46047 |
| Black/African/Caribbean/Black British | Other Black | 9t1D·00 | Black/Afri/Carib/Black Brit: other – NI eth cat 2011 census | 111880 |
| Black/African/Caribbean/Black British | Other Black | 9t0F·00 | Black/Afr/Carib/Black Brit: other Black– Eng+Wales 2011 cens | 110540 |
| Black/African/Caribbean/Black British | Other Black | 9t2G·00 | Carib/Black: any other Black/Caribbean grp – Scotland 2011 | 112216 |
| Black/African/Caribbean/Black British | Other Black | 9t2F·00 | Carib/Black: Black/Black Scot/Black Brit– Scotland 2011 cens | 112649 |
| Other ethnic group | Arab | 9S43·00 | Black N African/Arab/Iranian | 41329 |
| Other ethnic group | Arab | 9S43·12 | Black Arab | 57752 |
| Other ethnic group | Arab | 9SA4·00 | N African Arab/Iranian (NMO) | 24962 |
| Other ethnic group | Arab | 9SA4·11 | North African Arab (NMO) | 47285 |
| Other ethnic group | Arab | 9SA4·12 | Iranian (NMO) | 25082 |
| Other ethnic group | Arab | 9iF9·00 | Arab – ethnic category 2001 census | 46059 |
| Other ethnic group | Arab | 9t1E·00 | Other ethnic group: Arab – NI ethnic category 2011 census | 110780 |
| Other ethnic group | Arab | 9t0G·00 | Other ethnic group: Arab – Eng+Wales ethnic cat 2011 census | 110555 |
| Other ethnic group | Arab | 9t2H·00 | Other ethnic grp: Arab/Arab Scot/Arab British– Scotland 2011 | 112245 |
| Other ethnic group | Arab | 134C·00 | RACE: Arab | 12875 |
| Other ethnic group | Any other ethnic group | 9T1··00 | New Zealand ethnic groups | 45008 |
| Other ethnic group | Any other ethnic group | 9T1Y·00 | Other New Zealand ethnic group | 96789 |
| Other ethnic group | Any other ethnic group | 9T1Z·00 | New Zealand ethnic group NOS | 71425 |
| Other ethnic group | Any other ethnic group | 9SA··00 | Other ethnic non–mixed (NMO) | 30280 |
| Other ethnic group | Any other ethnic group | 9SA1·00 | Brit· ethnic minor· spec·(NMO) | 32110 |
| Other ethnic group | Any other ethnic group | 9SA2·00 | Brit· ethnic minor· unsp (NMO) | 57764 |
| Other ethnic group | Any other ethnic group | 9SAD·00 | Other ethnic NEC (NMO) | 41214 |
| Other ethnic group | Any other ethnic group | 9SJ··00 | Other ethnic group | 12757 |
| Other ethnic group | Any other ethnic group | 9T1A·00 | Other Pacific ethnic group | 46752 |
| Other ethnic group | Any other ethnic group | 9iF··00 | Other – ethnic category 2001 census | 12434 |
| Other ethnic group | Any other ethnic group | 9iF4·00 | Buddhist – ethnic category 2001 census | 63872 |
| Other ethnic group | Any other ethnic group | 9iF5·00 | Hindu – ethnic category 2001 census | 56127 |
| Other ethnic group | Any other ethnic group | 9iF6·00 | Jewish – ethnic category 2001 census | 46063 |
| Other ethnic group | Any other ethnic group | 9iF7·00 | Muslim – ethnic category 2001 census | 47091 |
| Other ethnic group | Any other ethnic group | 9iF8·00 | Sikh – ethnic category 2001 census | 49658 |
| Other ethnic group | Any other ethnic group | 9iFB·00 | Mid East (excl Israeli, Iranian & Arab) – eth cat 2001 cens | 28909 |
| Other ethnic group | Any other ethnic group | 9iFC·00 | Israeli – ethnic category 2001 census | 46964 |
| Other ethnic group | Any other ethnic group | 9iFD·00 | Iranian – ethnic category 2001 census | 25937 |
| Other ethnic group | Any other ethnic group | 9iFE·00 | Kurdish – ethnic category 2001 census | 45964 |
| Other ethnic group | Any other ethnic group | 9iFF·00 | Moroccan – ethnic category 2001 census | 25451 |
| Other ethnic group | Any other ethnic group | 9iFG·00 | Latin American – ethnic category 2001 census | 26246 |
| Other ethnic group | Any other ethnic group | 9iFH·00 | South and Central American – ethnic category 2001 census | 12756 |
| Other ethnic group | Any other ethnic group | 9iFJ·00 | Mauritian/Seychellois/Maldivian/St Helena eth cat 2001census | 32382 |
| Other ethnic group | Any other ethnic group | 9iFK·00 | Any other group – ethnic category 2001 census | 26455 |
| Other ethnic group | Any other ethnic group | 9t0H·00 | Other ethnic: any other grp – Eng+Wales eth cat 2011 census | 110742 |
| Other ethnic group | Any other ethnic group | 9t2J·00 | Other ethnic grp: any other ethnic grp– Scotland 2011 census | 111806 |
| Other ethnic group | Any other ethnic group | 9t1F·00 | Other ethnic group: any other grp– NI ethnic cat 2011 census | 110646 |
| Other ethnic group | Any other ethnic group | 9T11·11 | Pakeha | 85509 |
| Other ethnic group | Any other ethnic group | 9T13·00 | New Zealand Maori | 32479 |
| Other ethnic group | Any other ethnic group | 9T14·00 | Samoan | 64610 |
| Other ethnic group | Any other ethnic group | 9T15·00 | Cook Island Maori | 89910 |
| Other ethnic group | Any other ethnic group | 9T16·00 | Tongan | 60837 |
| Other ethnic group | Any other ethnic group | 9T17·00 | Niuean | 55584 |
| Other ethnic group | Any other ethnic group | 9T18·00 | Tokelauan | 25434 |
| Other ethnic group | Any other ethnic group | 9T19·00 | Fijian | 64609 |
| Other ethnic group | Any other ethnic group | 9T3··00 | Yemeni | 94487 |
| Other ethnic group | Any other ethnic group | 134P·11 | RACE: Other | 32391 |
| N/A | Ethnic group not specified | 916E·00 | Patient ethnicity unknown | 93749 |
| N/A | Ethnic group not specified | 9S···00 | Ethnic groups (1991 census) | 10196 |
| N/A | Ethnic group not specified | 9SD··00 | Ethnic group not given – patient refused | 12429 |
| N/A | Ethnic group not specified | 9SE··00 | Ethnic group not recorded | 24340 |
| N/A | Ethnic group not specified | 9SZ··00 | Ethnic groups (census) NOS | 45199 |
| N/A | Ethnic group not specified | 9T···00 | Ethnicity and other related nationality data | 23955 |
| N/A | Ethnic group not specified | 9i···00 | Ethnic category – 2001 census | 12435 |
| N/A | Ethnic group not specified | 9iG··00 | Ethnic category not stated – 2001 census | 12459 |
| N/A | Ethnic group not specified | 9t1··00 | Ethnic category – 2011 census Northern Ireland | 112302 |
| N/A | Ethnic group not specified | 9t0··00 | Ethnic category – 2011 census England and Wales | 110417 |
| N/A | Ethnic group not specified | 9t···00 | Ethnic category – 2011 census | 110472 |
| N/A | Ethnic group not specified | 9t2··00 | Ethnic category – 2011 census Scotland | 110962 |
| N/A | Ethnic group not specified | 134O·00 | RACE: Unknown | 22953 |
| N/A | Ethnic group not specified | 134P·00 | RACE: Not stated | 46137 |

Where a patient had multiple or conflicting ethnicity codes, their most recently recorded ethnicity was used. Where the date of ethnicity recording was unknown, the most frequently recorded ethnicity was used.

### Table S3: Demographic characteristics of the annual cohort

| **Characteristic** | **Overall**  N = 601,033 | **Non–migrant**  n = 455800 (75·8%) | **Migrant**  n = 145233 (24·2%) | **Definite**  n = 53729 (37·0%) | **Probable**  n=91504 (63·0%) |
| --- | --- | --- | --- | --- | --- |
| *Follow up, person–years* |  |  |  |  |  |
| Total | 1,353,914 | 1,054,984 | 298,930 | 103,743 | 195,188 |
| Mean (SD) | 2·25 (1·91) | 2·31 (1·93) | 2·06 (1·84) | 1·93 (1·82) | 2·13 (1·84) |
| Median (IQR) | 1·59 (2·98) | 1·61 (3·10) | 1·55 (2·73) | 1·44 (2·71) | 1·64 (2·75) |
| *Sex, n (%)* |  |  |  |  |  |
| Male | 294,377 (49·0%) | 225,228 (49·4%) | 69,149 (47·6%) | 25,881 (48·2%) | 43,268 (47·3%) |
| Female | 306,656 (51·0%) | 230,572 (50·6%) | 76,084 (52·4%) | 27,848 (51·8%) | 48,236 (52·7%) |
| *Year of cohort entry, n (%)* |  |  |  |  |  |
| 2015 | 488,966 (81·4%) | 389,449 (85·4%) | 99,517 (68·5%) | 42,228 (78·6%) | 57,289 (62·6%) |
| 2016 | 31,809 (5·3%) | 19,832 (4·4%) | 11,977 (8·2%) | 3,870 (7·2%) | 8,107 (8·9%) |
| 2017 | 27,971 (4·7%) | 18,005 (4·0%) | 9,966 (6·9%) | 2,637 (4·9%) | 7,329 (8·0%) |
| 2018 | 23,249 (3·9%) | 12,971 (2·8%) | 10,278 (7·1%) | 2,353 (4·4%) | 7,925 (8·7%) |
| 2019 | 25,380 (4·2%) | 12,478 (2·7%) | 12,902 (8·9%) | 2,519 (4·7%) | 10,383 (11·3%) |
| 2020 | 3,658 (0·6%) | 3,065 (0·7%) | 593 (0·4%) | 122 (0·2%) | 471 (0·5%) |
| *Age at cohort entry, years* |  |  |  |  |  |
| Mean (SD) | 38 (23) | 40 (24) | 33 (17) | 35 (16) | 32 (18) |
| Median (IQR) | 36 (33) | 39 (38) | 33 (19) | 33 (18) | 32 (21) |
| *Age at cohort exit, years* |  |  |  |  |  |
| Mean (SD) | 40 (23) | 41 (24) | 35 (17) | 36 (16) | 34 (18) |
| Median (IQR) | 38 (34) | 41 (38) | 34 (20) | 35 (19) | 34 (21) |
| *Time from database entry to cohort entry, years* |  |  |  |  |  |
| Mean (SD) | 7 (7) | 8 (7) | 2 (4) | 2·5 (3·7) | 2·3 (3·7) |
| Median (IQR) | 4 (13) | 7 (14) | 0 (3) | 0·5 (3·6) | 0·3 (3·4) |
| *Ethnicity, n (%)* |  |  |  |  |  |
| White British | 217,139 (36·1%) | 212,643 (46·7%) | 4,496 (3·1%) | 2,083 (3·9%) | 2,413 (2·6%) |
| White non–British | 89,122 (14·8%) | 33,343 (7·3%) | 55,779 (38·4%) | 15,299 (28·5%) | 40,480 (44·2%) |
| Mixed/Multiple ethnic groups | 9,345 (1·6%) | 5,117 (1·1%) | 4,228 (2·9%) | 1,736 (3·2%) | 2,492 (2·7%) |
| Asian/Asian British | 55,265 (9·2%) | 15,414 (3·4%) | 39,851 (27·4%) | 11,412 (21·2%) | 28,439 (31·1%) |
| Black/African/Caribbean/Black British | 25,084 (4·2%) | 11,529 (2·5%) | 13,555 (9·3%) | 7,230 (13·5%) | 6,325 (6·9%) |
| Other ethnic group | 12,842 (2·1%) | 3,028 (0·7%) | 9,814 (6·8%) | 2,962 (5·5%) | 6,852 (7·5%) |
| Unknown | 192,236 (32·0%) | 174,726 (38·3%) | 17,510 (12·1%) | 13,007 (24·2%) | 4,503 (4·9%) |
| *Practice region, n (%)* |  |  |  |  |  |
| London | 136,726 (22·7%) | 74,966 (16·4%) | 61,760 (42·5%) | 28,369 (52·8%) | 33,391 (36·5%) |
| North East | 6,869 (1·1%) | 6,373 (1·4%) | 496 (0·3%) | 77 (0·1%) | 419 (0·5%) |
| North West | 80,019 (13·3%) | 65,528 (14·4%) | 14,491 (10·0%) | 4,769 (8·9%) | 9,722 (10·6%) |
| Yorkshire & The Humber | 8,519 (1·4%) | 8,207 (1·8%) | 312 (0·2%) | 74 (0·1%) | 238 (0·3%) |
| East Midlands | 674 (0·1%) | 628 (0·1%) | 46 (0·0%) | 14 (0·0%) | 32 (0·0%) |
| West Midlands | 61,639 (10·3%) | 51,596 (11·3%) | 10,043 (6·9%) | 1,333 (2·5%) | 8,710 (9·5%) |
| East of England | 45,622 (7·6%) | 37,426 (8·2%) | 8,196 (5·6%) | 998 (1·9%) | 7,198 (7·9%) |
| South West | 56,161 (9·3%) | 47,956 (10·5%) | 8,205 (5·6%) | 3,727 (6·9%) | 4,478 (4·9%) |
| South Central | 93,368 (15·5%) | 72,188 (15·8%) | 21,180 (14·6%) | 10,889 (20·3%) | 10,291 (11·2%) |
| South East Coast | 111,436 (18·5%) | 90,932 (19·9%) | 20,504 (14·1%) | 3,479 (6·5%) | 17,025 (18·6%) |
| *IMD, n (%)* |  |  |  |  |  |
| IMD 1 | 132,635 (22·1%) | 114,157 (25·0%) | 18,478 (12·7%) | 7,768 (14·5%) | 10,710 (11·7%) |
| IMD 2 | 115,950 (19·3%) | 96,212 (21·1%) | 19,738 (13·6%) | 6,206 (11·6%) | 13,532 (14·8%) |
| IMD 3 | 117,655 (19·6%) | 91,493 (20·1%) | 26,162 (18·0%) | 8,561 (15·9%) | 17,601 (19·2%) |
| IMD 4 | 122,409 (20·4%) | 82,710 (18·1%) | 39,699 (27·3%) | 14,835 (27·6%) | 24,864 (27·2%) |
| IMD 5 | 112,384 (18·7%) | 71,228 (15·6%) | 41,156 (28·3%) | 16,359 (30·4%) | 24,797 (27·1%) |
| *Patients in each study year, n (%)* |  |  |  |  |  |
| 2015 | 488,966 (29·2%) | 389,449 (30·2%) | 99,517 (25·8%) | 42,228 (31·5%) | 57,289 (22·7%) |
| 2016 | 350,071 (20·9%) | 277,077 (21·5%) | 72,994 (18·9%) | 28,538 (21·3%) | 44,456 (17·6%) |
| 2017 | 271,309 (16·2%) | 210,582 (16·3%) | 60,727 (15·7%) | 19,255 (14·4%) | 41,472 (16·5%) |
| 2018 | 226,471 (13·5%) | 167,070 (13·0%) | 59,401 (15·4%) | 18,604 (13·9%) | 40,797 (16·2%) |
| 2019 | 200,161 (12·0%) | 144,785 (11·2%) | 55,376 (14·3%) | 14,857 (11·1%) | 40,519 (16·1%) |
| 2020 | 137,410 (8·2%) | 99,248 (7·7%) | 38,162 (9·9%) | 10,658 (7·9%) | 27,504 (10·9%) |

### Table S4: Demographic characteristics of the annual cohort matched on age at and year of contributing up–to–standard data to the CPRD GOLD database and practice region

| **Characteristic** | **Overall**  **N = 244,266** | **Non–migrant**  **n = 122133 (50·0%)** | **Migrant**  **n = 122133 (50·0%)** | **Definite**  **n = 41593 (34·1%)** | **Probable**  **n = 80540 (65·9%)** |
| --- | --- | --- | --- | --- | --- |
| *Follow up, person–years* |  |  |  |  |  |
| Total | 524,732 | 259,103 | 265,630 | 88,042 | 177,588 |
| Mean (SD) | 2·15 (1·87) | 2·12 (1·89) | 2·17 (1·86) | 2·12 (1·86) | 2·20 (1·85) |
| Median (IQR) | 1·59 (2·86) | 1·52 (2·91) | 1·71 (2·82) | 1·62 (2·87) | 1·75 (2·81) |
| *Sex, n (%)* |  |  |  |  |  |
| Male | 118,204 (48·4%) | 60,037 (49·2%) | 58,167 (47·6%) | 20,143 (48·4%) | 38,024 (47·2%) |
| Female | 126,062 (51·6%) | 62,096 (50·8%) | 63,966 (52·4%) | 21,450 (51·6%) | 42,516 (52·8%) |
| *Year of cohort entry, n (%)* |  |  |  |  |  |
| 2015 | 169,746 (69·5%) | 84,873 (69·5%) | 84,873 (69·5%) | 32,199 (77·4%) | 52,674 (65·4%) |
| 2016 | 21,132 (8·7%) | 10,566 (8·7%) | 10,566 (8·7%) | 3,333 (8·0%) | 7,233 (9·0%) |
| 2017 | 18,064 (7·4%) | 9,032 (7·4%) | 9,032 (7·4%) | 2,417 (5·8%) | 6,615 (8·2%) |
| 2018 | 16,208 (6·6%) | 8,104 (6·6%) | 8,104 (6·6%) | 1,799 (4·3%) | 6,305 (7·8%) |
| 2019 | 18,048 (7·4%) | 9,024 (7·4%) | 9,024 (7·4%) | 1,739 (4·2%) | 7,285 (9·0%) |
| 2020 | 1,068 (0·4%) | 534 (0·4%) | 534 (0·4%) | 106 (0·3%) | 428 (0·5%) |
| *Age at cohort entry, years* |  |  |  |  |  |
| Mean (SD) | 33 (18) | 33 (18) | 33 (18) | 35 (17) | 32 (19) |
| Median (IQR) | 33 (22) | 33 (22) | 33 (22) | 34 (21) | 32 (24) |
| *Age at cohort exit, years* |  |  |  |  |  |
| Mean (SD) | 35 (18) | 35 (18) | 35 (18) | 37 (17) | 34 (19) |
| Median (IQR) | 34 (23) | 34 (23) | 35 (23) | 36 (21) | 34 (24) |
| *Time between CPRD GOLD entry to study entry, years* |  |  |  |  |  |
| Mean (SD) | 2·6 (3·9) | 2·6 (3·9) | 2·6 (3·9) | 2·9 (3·9) | 2·5 (3·9) |
| Median (IQR) | 0·8 (4·0) | 0·8 (4·0) | 0·8 (4·0) | 0·9 (4·6) | 0·7 (3·7) |
| *Ethnicity, n (%)* |  |  |  |  |  |
| White British | 64,027 (26·2%) | 60,193 (49·3%) | 3,834 (3·1%) | 1,660 (4·0%) | 2,174 (2·7%) |
| White non–British | 59,889 (24·5%) | 12,490 (10·2%) | 47,399 (38·8%) | 11,785 (28·3%) | 35,614 (44·2%) |
| Mixed/Multiple ethnic groups | 6,026 (2·5%) | 2,440 (2·0%) | 3,586 (2·9%) | 1,359 (3·3%) | 2,227 (2·8%) |
| Asian/Asian British | 41,445 (17·0%) | 7,701 (6·3%) | 33,744 (27·6%) | 8,807 (21·2%) | 24,937 (31·0%) |
| Black/African/Caribbean/Black British | 18,414 (7·5%) | 6,863 (5·6%) | 11,551 (9·5%) | 5,938 (14·3%) | 5,613 (7·0%) |
| Other ethnic group | 10,215 (4·2%) | 1,641 (1·3%) | 8,574 (7·0%) | 2,504 (6·0%) | 6,070 (7·5%) |
| Unknown | 44,250 (18·1%) | 30,805 (25·2%) | 13,445 (11·0%) | 9,540 (22·9%) | 3,905 (4·8%) |
| *Practice region, n (%)* |  |  |  |  |  |
| London | 95,674 (39·2%) | 47,837 (39·2%) | 47,837 (39·2%) | 21,772 (52·3%) | 26,065 (32·4%) |
| North East | 846 (0·3%) | 423 (0·3%) | 423 (0·3%) | 65 (0·2%) | 358 (0·4%) |
| North West | 27,356 (11·2%) | 13,678 (11·2%) | 13,678 (11·2%) | 4,546 (10·9%) | 9,132 (11·3%) |
| Yorkshire & The Humber | 558 (0·2%) | 279 (0·2%) | 279 (0·2%) | 58 (0·1%) | 221 (0·3%) |
| East Midlands | 62 (0·0%) | 31 (0·0%) | 31 (0·0%) | 8 (0·0%) | 23 (0·0%) |
| West Midlands | 19,460 (8·0%) | 9,730 (8·0%) | 9,730 (8·0%) | 1,275 (3·1%) | 8,455 (10·5%) |
| East of England | 13,534 (5·5%) | 6,767 (5·5%) | 6,767 (5·5%) | 908 (2·2%) | 5,859 (7·3%) |
| South West | 15,572 (6·4%) | 7,786 (6·4%) | 7,786 (6·4%) | 3,555 (8·5%) | 4,231 (5·3%) |
| South Central | 30,656 (12·6%) | 15,328 (12·6%) | 15,328 (12·6%) | 5,969 (14·4%) | 9,359 (11·6%) |
| South East Coast | 40,548 (16·6%) | 20,274 (16·6%) | 20,274 (16·6%) | 3,437 (8·3%) | 16,837 (20·9%) |
| *IMD, n (%)* |  |  |  |  |  |
| IMD 1 | 43,400 (17·8%) | 28,537 (23·4%) | 14,863 (12·2%) | 5,297 (12·7%) | 9,566 (11·9%) |
| IMD 2 | 41,592 (17·0%) | 24,748 (20·3%) | 16,844 (13·8%) | 4,896 (11·8%) | 11,948 (14·8%) |
| IMD 3 | 44,623 (18·3%) | 23,280 (19·1%) | 21,343 (17·5%) | 6,112 (14·7%) | 15,231 (18·9%) |
| IMD 4 | 57,071 (23·4%) | 24,046 (19·7%) | 33,025 (27·0%) | 11,483 (27·6%) | 21,542 (26·7%) |
| IMD 5 | 57,580 (23·6%) | 21,522 (17·6%) | 36,058 (29·5%) | 13,805 (33·2%) | 22,253 (27·6%) |
| *Patients in each study year, n (%)* |  |  |  |  |  |
| 2015 | 169,746 (25·3%) | 84,873 (25·5%) | 84,873 (25·2%) | 32,199 (29·0%) | 52,674 (23·3%) |
| 2016 | 127,934 (19·1%) | 62,780 (18·8%) | 65,154 (19·3%) | 24,158 (21·8%) | 40,996 (18·1%) |
| 2017 | 110,600 (16·5%) | 55,760 (16·7%) | 54,840 (16·3%) | 16,841 (15·2%) | 37,999 (16·8%) |
| 2018 | 102,080 (15·2%) | 49,771 (14·9%) | 52,309 (15·5%) | 15,958 (14·4%) | 36,351 (16·1%) |
| 2019 | 95,147 (14·2%) | 48,327 (14·5%) | 46,820 (13·9%) | 12,539 (11·3%) | 34,281 (15·1%) |
| 2020 | 64,960 (9·7%) | 31,643 (9·5%) | 33,317 (9·9%) | 9,246 (8·3%) | 24,071 (10·6%) |

### Table S5: Demographic characteristics of the annual cohort matched on follow–up time (person–years) and practice region

| **Characteristic** | **Overall**  N = 233,894 | **Non–migrant**  n = 116947 (50·0%) | **Migrant**  n = 116947 (50·0%) | **Definite**  n = 37620 (32·2%) | **Probable**  n = 79327 (67·8%) |
| --- | --- | --- | --- | --- | --- |
| *Follow up, person–years* |  |  |  |  |  |
| Total | 487,038 | 243,519 | 243,519 | 77,605 | 165,914 |
| Mean (SD) | 2·08 (1·85) | 2·08 (1·85) | 2·08 (1·85) | 2·06 (1·89) | 2·09 (1·84) |
| Median (IQR) | 1·51 (2·84) | 1·51 (2·84) | 1·51 (2·84) | 1·46 (2·88) | 1·53 (2·71) |
| *Sex, n (%)* |  |  |  |  |  |
| Male | 113,058 (48·3%) | 57,910 (49·5%) | 55,148 (47·2%) | 17,854 (47·5%) | 37,294 (47·0%) |
| Female | 120,836 (51·7%) | 59,037 (50·5%) | 61,799 (52·8%) | 19,766 (52·5%) | 42,033 (53·0%) |
| *Year of cohort entry, n (%)* |  |  |  |  |  |
| 2015 | 162,087 (69·3%) | 84,388 (72·2%) | 77,699 (66·4%) | 27,815 (73·9%) | 49,884 (62·9%) |
| 2016 | 18,084 (7·7%) | 8,038 (6·9%) | 10,046 (8·6%) | 3,168 (8·4%) | 6,878 (8·7%) |
| 2017 | 16,499 (7·1%) | 7,909 (6·8%) | 8,590 (7·3%) | 2,187 (5·8%) | 6,403 (8·1%) |
| 2018 | 15,631 (6·7%) | 6,888 (5·9%) | 8,743 (7·5%) | 1,974 (5·2%) | 6,769 (8·5%) |
| 2019 | 19,388 (8·3%) | 8,088 (6·9%) | 11,300 (9·7%) | 2,359 (6·3%) | 8,941 (11·3%) |
| 2020 | 2,205 (0·9%) | 1,636 (1·4%) | 569 (0·5%) | 117 (0·3%) | 452 (0·6%) |
| *Age at cohort entry, years* |  |  |  |  |  |
| Mean (SD) | 35 (21) | 37 (24) | 33 (17) | 35 (16) | 32 (18) |
| Median (IQR) | 33 (27) | 34 (36) | 33 (20) | 34 (18) | 32 (21) |
| *Age at cohort exit, years* |  |  |  |  |  |
| Mean (SD) | 37 (21) | 38 (24) | 35 (17) | 37 (16) | 34 (18) |
| Median (IQR) | 35 (27) | 36 (37) | 35 (20) | 36 (19) | 34 (21) |
| *Time from CPRD GOLD entry to study entry, years* |  |  |  |  |  |
| Mean (SD) | 3·9 (5·7) | 5·5 (6·8) | 2·4 (3·7) | 2·6 (3·8) | 2·3 (3·7) |
| Median (IQR) | 1·1 (5·7) | 2·2 (10·4) | 0·5 (3·6) | 0·6 (4·0) | 0·4 (3·4) |
| *Ethnicity, n (%)* |  |  |  |  |  |
| White British | 58,593 (25·1%) | 54,866 (46·9%) | 3,727 (3·2%) | 1,580 (4·2%) | 2,147 (2·7%) |
| White non–British | 56,962 (24·4%) | 10,291 (8·8%) | 46,671 (39·9%) | 11,051 (29·4%) | 35,620 (44·9%) |
| Mixed/Multiple ethnic groups | 5,736 (2·5%) | 2,294 (2·0%) | 3,442 (2·9%) | 1,232 (3·3%) | 2,210 (2·8%) |
| Asian/Asian British | 39,546 (16·9%) | 7,385 (6·3%) | 32,161 (27·5%) | 7,824 (20·8%) | 24,337 (30·7%) |
| Black/African/Caribbean/Black British | 16,333 (7·0%) | 5,832 (5·0%) | 10,501 (9·0%) | 5,116 (13·6%) | 5,385 (6·8%) |
| Other ethnic group | 9,452 (4·0%) | 1,405 (1·2%) | 8,047 (6·9%) | 2,298 (6·1%) | 5,749 (7·2%) |
| Unknown | 47,272 (20·2%) | 34,874 (29·8%) | 12,398 (10·6%) | 8,519 (22·6%) | 3,879 (4·9%) |
| *Practice region, n (%)* |  |  |  |  |  |
| London | 89,240 (38·2%) | 44,620 (38·2%) | 44,620 (38·2%) | 18,753 (49·8%) | 25,867 (32·6%) |
| North East | 692 (0·3%) | 346 (0·3%) | 346 (0·3%) | 62 (0·2%) | 284 (0·4%) |
| North West | 26,982 (11·5%) | 13,491 (11·5%) | 13,491 (11·5%) | 4,428 (11·8%) | 9,063 (11·4%) |
| Yorkshire & The Humber | 528 (0·2%) | 264 (0·2%) | 264 (0·2%) | 65 (0·2%) | 199 (0·3%) |
| East Midlands | 88 (0·0%) | 44 (0·0%) | 44 (0·0%) | 14 (0·0%) | 30 (0·0%) |
| West Midlands | 18,724 (8·0%) | 9,362 (8·0%) | 9,362 (8·0%) | 1,195 (3·2%) | 8,167 (10·3%) |
| East of England | 14,614 (6·2%) | 7,307 (6·2%) | 7,307 (6·2%) | 936 (2·5%) | 6,371 (8·0%) |
| South West | 14,612 (6·2%) | 7,306 (6·2%) | 7,306 (6·2%) | 3,152 (8·4%) | 4,154 (5·2%) |
| South Central | 30,080 (12·9%) | 15,040 (12·9%) | 15,040 (12·9%) | 5,604 (14·9%) | 9,436 (11·9%) |
| South East Coast | 38,334 (16·4%) | 19,167 (16·4%) | 19,167 (16·4%) | 3,411 (9·1%) | 15,756 (19·9%) |
| *IMD, n (%)* |  |  |  |  |  |
| IMD 1 | 41,061 (17·6%) | 25,888 (22·1%) | 15,173 (13·0%) | 5,433 (14·4%) | 9,740 (12·3%) |
| IMD 2 | 40,249 (17·2%) | 23,499 (20·1%) | 16,750 (14·3%) | 4,706 (12·5%) | 12,044 (15·2%) |
| IMD 3 | 43,172 (18·5%) | 22,418 (19·2%) | 20,754 (17·7%) | 5,633 (15·0%) | 15,121 (19·1%) |
| IMD 4 | 53,261 (22·8%) | 22,815 (19·5%) | 30,446 (26·0%) | 9,745 (25·9%) | 20,701 (26·1%) |
| IMD 5 | 56,151 (24·0%) | 22,327 (19·1%) | 33,824 (28·9%) | 12,103 (32·2%) | 21,721 (27·4%) |
| *Patients in each study year, n (%)* |  |  |  |  |  |
| 2015 | 162,087 (25·8%) | 84,388 (27·0%) | 77,699 (24·7%) | 27,815 (27·9%) | 49,884 (23·2%) |
| 2016 | 119,649 (19·1%) | 61,142 (19·5%) | 58,507 (18·6%) | 20,402 (20·5%) | 38,105 (17·7%) |
| 2017 | 103,246 (16·4%) | 51,827 (16·6%) | 51,419 (16·3%) | 15,509 (15·6%) | 35,910 (16·7%) |
| 2018 | 95,521 (15·2%) | 45,902 (14·7%) | 49,619 (15·7%) | 14,871 (14·9%) | 34,748 (16·1%) |
| 2019 | 88,530 (14·1%) | 41,504 (13·3%) | 47,026 (14·9%) | 12,535 (12·6%) | 34,491 (16·0%) |
| 2020 | 58,924 (9·4%) | 28,111 (9·0%) | 30,813 (9·8%) | 8,554 (8·6%) | 22,259 (10·3%) |

### Table S6: Demographic characteristics of the interrupted time series cohort matched on age and year of joining the CPRD GOLD database, sex, practice region and IMD

| **Characteristic** | **Overall**  N = 192,336 | **Non–migrant**  n = 96168 (50·0%) | **Migrant**  n = 96168 (50·0%) | **Definite**  n = 32531 (33·8%) | **Probable**  n = 63637 (66·2%) |
| --- | --- | --- | --- | --- | --- |
| *Follow up, person–years* |  |  |  |  |  |
| Total | 413,441 | 201,533 | 211,908 | 68,392 | 143,515 |
| Mean (SD) | 2·15 (1·88) | 2·10 (1·88) | 2·20 (1·87) | 2·10 (1·87) | 2·26 (1·87) |
| Median (IQR) | 1·59 (2·86) | 1·49 (2·87) | 1·72 (2·85) | 1·59 (2·87) | 1·78 (2·86) |
| *Sex, n (%)* |  |  |  |  |  |
| Male | 91,588 (47·6%) | 45,794 (47·6%) | 45,794 (47·6%) | 15,802 (48·6%) | 29,992 (47·1%) |
| Female | 100,748 (52·4%) | 50,374 (52·4%) | 50,374 (52·4%) | 16,729 (51·4%) | 33,645 (52·9%) |
| *Year of cohort entry, n (%)* |  |  |  |  |  |
| 2015 | 137,640 (71·6%) | 68,820 (71·6%) | 68,820 (71·6%) | 26,124 (80·3%) | 42,696 (67·1%) |
| 2016 | 15,252 (7·9%) | 7,626 (7·9%) | 7,626 (7·9%) | 2,237 (6·9%) | 5,389 (8·5%) |
| 2017 | 13,582 (7·1%) | 6,791 (7·1%) | 6,791 (7·1%) | 1,682 (5·2%) | 5,109 (8·0%) |
| 2018 | 12,408 (6·5%) | 6,204 (6·5%) | 6,204 (6·5%) | 1,344 (4·1%) | 4,860 (7·6%) |
| 2019 | 12,630 (6·6%) | 6,315 (6·6%) | 6,315 (6·6%) | 1,073 (3·3%) | 5,242 (8·2%) |
| 2020 | 824 (0·4%) | 412 (0·4%) | 412 (0·4%) | 71 (0·2%) | 341 (0·5%) |
| *Age at cohort entry, years* |  |  |  |  |  |
| Mean (SD) | 32 (18) | 32 (18) | 32 (18) | 35 (16) | 31 (18) |
| Median (IQR) | 32 (22) | 32 (22) | 32 (22) | 34 (20) | 31 (24) |
| *Age at cohort exit, years* |  |  |  |  |  |
| Mean (SD) | 34 (18) | 34 (18) | 34 (18) | 36 (16) | 33 (18) |
| Median (IQR) | 34 (22) | 34 (23) | 34 (22) | 35 (21) | 33 (24) |
| *Time from CPRD GOLD entry to study entry, years* |  |  |  |  |  |
| Mean (SD) | 2·6 (3·9) | 2·6 (3·9) | 2·6 (3·9) | 2·8 (3·9) | 2·5 (3·9) |
| Median (IQR) | 0·8 (4·0) | 0·8 (4·0) | 0·8 (4·0) | 0·9 (4·5) | 0·8 (3·8) |
| *Ethnicity, n (%)* |  |  |  |  |  |
| White British | 48,696 (25·3%) | 45,418 (47·2%) | 3,278 (3·4%) | 1,439 (4·4%) | 1,839 (2·9%) |
| White non–British | 48,225 (25·1%) | 10,055 (10·5%) | 38,170 (39·7%) | 9,195 (28·3%) | 28,975 (45·5%) |
| Mixed/Multiple ethnic groups | 4,979 (2·6%) | 2,128 (2·2%) | 2,851 (3·0%) | 1,067 (3·3%) | 1,784 (2·8%) |
| Asian/Asian British | 32,308 (16·8%) | 6,567 (6·8%) | 25,741 (26·8%) | 6,811 (20·9%) | 18,930 (29·7%) |
| Black/African/Caribbean/Black British | 14,768 (7·7%) | 6,245 (6·5%) | 8,523 (8·9%) | 4,289 (13·2%) | 4,234 (6·7%) |
| Other ethnic group | 8,157 (4·2%) | 1,431 (1·5%) | 6,726 (7·0%) | 1,895 (5·8%) | 4,831 (7·6%) |
| Unknown | 35,203 (18·3%) | 24,324 (25·3%) | 10,879 (11·3%) | 7,835 (24·1%) | 3,044 (4·8%) |
| *Practice region, n (%)* |  |  |  |  |  |
| London | 75,916 (39·5%) | 37,958 (39·5%) | 37,958 (39·5%) | 16,683 (51·3%) | 21,275 (33·4%) |
| North East | 510 (0·3%) | 255 (0·3%) | 255 (0·3%) | 36 (0·1%) | 219 (0·3%) |
| North West | 21,268 (11·1%) | 10,634 (11·1%) | 10,634 (11·1%) | 3,449 (10·6%) | 7,185 (11·3%) |
| Yorkshire & The Humber | 248 (0·1%) | 124 (0·1%) | 124 (0·1%) | 16 (0·0%) | 108 (0·2%) |
| East Midlands | 42 (0·0%) | 21 (0·0%) | 21 (0·0%) | 4 (0·0%) | 17 (0·0%) |
| West Midlands | 14,250 (7·4%) | 7,125 (7·4%) | 7,125 (7·4%) | 889 (2·7%) | 6,236 (9·8%) |
| East of England | 8,544 (4·4%) | 4,272 (4·4%) | 4,272 (4·4%) | 723 (2·2%) | 3,549 (5·6%) |
| South West | 11,858 (6·2%) | 5,929 (6·2%) | 5,929 (6·2%) | 2,532 (7·8%) | 3,397 (5·3%) |
| South Central | 26,860 (14·0%) | 13,430 (14·0%) | 13,430 (14·0%) | 5,394 (16·6%) | 8,036 (12·6%) |
| South East Coast | 32,840 (17·1%) | 16,420 (17·1%) | 16,420 (17·1%) | 2,805 (8·6%) | 13,615 (21·4%) |
| *IMD, n (%)* |  |  |  |  |  |
| IMD 1 | 28,916 (15·0%) | 14,458 (15·0%) | 14,458 (15·0%) | 5,073 (15·6%) | 9,385 (14·7%) |
| IMD 2 | 31,276 (16·3%) | 15,638 (16·3%) | 15,638 (16·3%) | 4,588 (14·1%) | 11,050 (17·4%) |
| IMD 3 | 36,022 (18·7%) | 18,011 (18·7%) | 18,011 (18·7%) | 5,211 (16·0%) | 12,800 (20·1%) |
| IMD 4 | 47,740 (24·8%) | 23,870 (24·8%) | 23,870 (24·8%) | 8,172 (25·1%) | 15,698 (24·7%) |
| IMD 5 | 48,382 (25·2%) | 24,191 (25·2%) | 24,191 (25·2%) | 9,487 (29·2%) | 14,704 (23·1%) |
| *Patients in each study year, n (%)* |  |  |  |  |  |
| 2015 | 137,640 (26·1%) | 68,820 (26·5%) | 68,820 (25·7%) | 26,124 (30·3%) | 42,696 (23·5%) |
| 2016 | 101,227 (19·2%) | 49,673 (19·1%) | 51,554 (19·3%) | 18,611 (21·6%) | 32,943 (18·2%) |
| 2017 | 86,200 (16·4%) | 43,151 (16·6%) | 43,049 (16·1%) | 12,723 (14·8%) | 30,326 (16·7%) |
| 2018 | 78,646 (14·9%) | 37,831 (14·6%) | 40,815 (15·3%) | 12,085 (14·0%) | 28,730 (15·8%) |
| 2019 | 73,112 (13·9%) | 36,361 (14·0%) | 36,751 (13·7%) | 9,442 (11·0%) | 27,309 (15·0%) |
| 2020 | 50,295 (9·5%) | 23,672 (9·1%) | 26,623 (9·9%) | 7,135 (8·3%) | 19,488 (10·7%) |

### Table S7: Crude consultations rates 2015–2020 by migration status and migration certainty in England

| **Year** | **Crude consultation rate per person–year (95%CI)** | | | |
| --- | --- | --- | --- | --- |
|  | **Non–migrant** | **Migrant** | **Definite** | **Probable** |
| 2015–2019 | 5·62 (5·62–5·62) | 4·31 (4·31–4·32) | 4·62 (4·61–4·64) | 4·14 (4·13–4·15) |
| 2015 | 5·66 (5·65–5·67) | 4·29 (4·28–4·31) | 4·06 (4·04–4·09) | 4·45 (4·43–4·47) |
| 2016 | 5·71 (5·7–5·72) | 4·41 (4·39–4·42) | 4·54 (4·51–4·57) | 4·33 (4·3–4·35) |
| 2017 | 5·59 (5·58–5·6) | 4·46 (4·45–4·48) | 5·14 (5·11–5·18) | 4·13 (4·11–4·15) |
| 2018 | 5·53 (5·51–5·54) | 4·19 (4·17–4·21) | 5·01 (4·98–5·05) | 3·8 (3·78–3·83) |
| 2019 | 5·5 (5·48–5·51) | 4·17 (4·15–4·19) | 5 (4·96–5·05) | 3·84 (3·82–3·86) |
| 2020 | 4·95 (4·94–4·97) | 3·99 (3·97–4·01) | 4·84 (4·79–4·88) | 3·65 (3·63–3·68) |

### Table S8: Crude consultations rates before and during the pandemic by migration status and migration certainty in England

|  |  |  | **Crude consultation rate per person–year (95%CI)** | | | |
| --- | --- | --- | --- | --- | --- | --- |
| Consultation type | Pandemic |  | Non–migrant | Migrant | Definite | Probable |
| All | Before |  | 4·6 (4·59–4·6) | 4·35 (4·34–4·36) | 4·73 (4·71–4·74) | 4·16 (4·15–4·17) |
|  | During |  | 4·2 (4·17–4·23) | 3·54 (3·52–3·57) | 4·33 (4·27–4·38) | 3·24 (3·21–3·27) |
| Face–to–face | Before |  | 4·35 (4·34–4·36) | 4·12 (4·11–4·13) | 4·48 (4·47–4·5) | 3·94 (3·93–3·95) |
|  | During |  | 3·52 (3·49–3·55) | 3·02 (3–3·05) | 3·77 (3·72–3·82) | 2·73 (2·7–2·76) |
| Telephone | Before |  | 0·25 (0·25–0·25) | 0·23 (0·23–0·23) | 0·24 (0·24–0·25) | 0·22 (0·22–0·22) |
|  | During |  | 0·68 (0·67–0·69) | 0·52 (0·51–0·53) | 0·56 (0·54–0·58) | 0·51 (0·5–0·52) |

### Table S9: Consultation rate ratios from interrupted time–series analysis (5 January 2015 to 26 December 2020) in England by age group

| **Variable** | **RR (95%CI)** | | | | | |
| --- | --- | --- | --- | --- | --- | --- |
|  | **0–15 years** | **16–24 years** | **25–34 years** | **35–49 years** | **50–64 years** | **65 and over** |
| Pandemic | 0·84  (0·79–0·89) | 0·92  (0·87–0·97) | 0·99  (0·94–1·04) | 0·99  (0·95–1·04) | 0·95  (0·9–1) | 0·91  (0·86–0·96) |
| Migration status | 1·05  (1·02–1·07) | 0·69  (0·68–0·71) | 0·74  (0·72–0·75) | 0·9  (0·88–0·92) | 1·13  (1·11–1·16) | 1·21  (1·18–1·23) |
| Migration status +  interaction term (between migrant status and pandemic) | 0·76  (0·71–0·82) | 0·59  (0·55–0·64) | 0·58  (0·55–0·62) | 0·74  (0·7–0·78) | 1·08  (1·02–1·15) | 1·28  (1·2–1·36) |
| Interaction term (between migrant status and pandemic) | 0·73  (0·68–0·79) | 0·86  (0·8–0·92) | 0·79  (0·74–0·84) | 0·82  (0·77–0·87) | 0·96  (0·9–1·02) | 1·06  (0·99–1·13) |

### Figure S1: Weekly consultation rates by migration status in London: predicted rates from interrupted time–series analysis (solid line) and actual observed rates (dashed line), truncated view July 2019 – November 2020

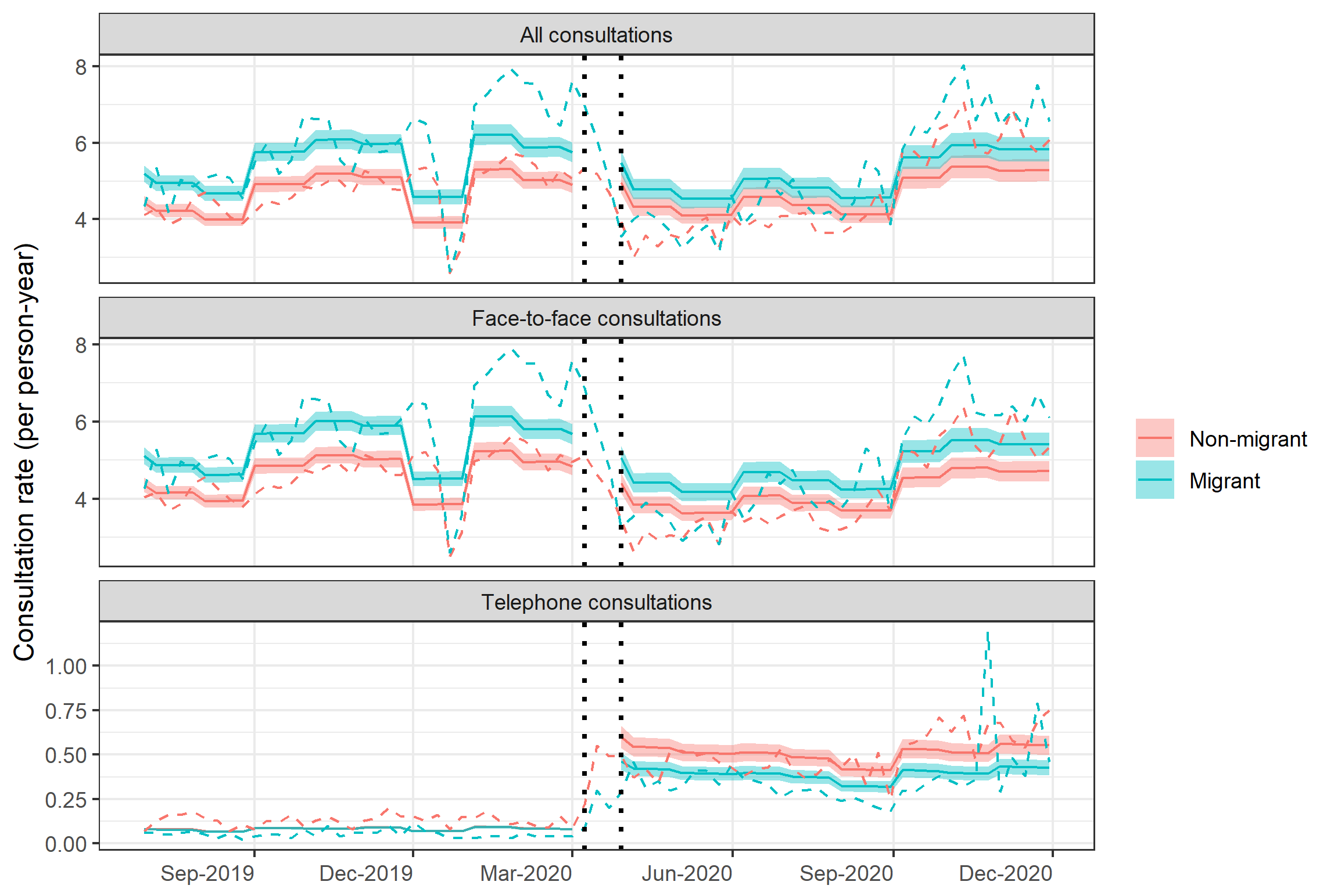

### Table S10: Consultation rate ratios from interrupted time series analysis (5 January 2015 to 26 December 2020) for London

| Variable | **Interpretation** | **All** | **Face–to–face** | **Telephone** |
| --- | --- | --- | --- | --- |
| Pandemic | Comparing non–migrants’ consultation rates during the pandemic to non–migrants’ consultation rates pre–pandemic | 1·01  (0·96–1·06) | 0·91  (0·86–0·96) | 7·6  (6·92–8·34) |
| Migration status | Comparing migrants’ consultation rates pre–pandemic to non–migrants consultations pre–pandemic | 1·17  (1·15–1·2) | 1·17  (1·15–1·2) | 0·99  (0·95–1·03) |
| Migration status +  interaction term (between migrant status and pandemic) | Comparing migrants’ consultation rates during the pandemic to non–migrants’ consultation rates during the pandemic | 1·1  (1·04–1·17) | 1·15  (1·08–1·22) | 0·77  (0·7–0·86) |
| Interaction term (between migrant status and pandemic) | Comparing the difference in consultation rates between migrants and non–migrants pre–pandemic to the difference in consultation rates between migrants and non–migrants during the pandemic (the additional effect of the pandemic on the difference between migrants and non–migrants i·e· the multiplicative effect) | 0·94  (0·88–1·01) | 0·98  (0·92–1·05) | 0·78  (0·7–0·88) |

### Table S11: Effect modification by ethnicity for the effect of migration on pre–pandemic consultation rates

|  | **RR (95% CI) comparing each group to White British non–migrants** | | **RR (95% CI) comparing migrants to non–migrants of each ethnicity** | | **Multiplicative effect (95%CI)** | **RERI, additive effect (95%CI)** | |
| --- | --- | --- | --- | --- | --- | --- | --- |
| Ethnicity | Non–migrants | Migrants |  |  |  |  |  |
| White British | 1·0 | 0·95 (0·92–0·97) | 0·95 (0·92–0·97) | | – | – | |
| White non–British | 0·92 (0·91–0·92) | 0·69 (0·68–0·69) | 0·75 (0·74–0·76) | | 0·79 (0·77–0·82) | –0·17 (–0·2––0·15) | |
| Mixed/Multiple ethnic groups | 0·96 (0·94–0·98) | 0·86 (0·84–0·88) | 0·9 (0·87–0·93) | | 0·95 (0·91–0·99) | –0·05 (–0·08––0·01) | |
| Asian/Asian British | 0·99 (0·97–1) | 1·17 (1·16–1·18) | 1·19 (1·17–1·21) | | 1·26 (1·22–1·29) | 0·24 (0·21–0·27) | |
| Black/African/ Caribbean/Black British | 0·88 (0·87–0·9) | 0·99 (0·98–1·01) | 1·13 (1·1–1·15) | | 1·19 (1·15–1·23) | 0·17 (0·14–0·2) | |
| Other ethnic group | 0·89 (0·87–0·92) | 0·91 (0·89–0·92) | 1·02 (0·98–1·05) | | 1·07 (1·03–1·12) | 0·07 (0·03–0·11) | |
| Unknown | 0·85 (0·85–0·86) | 0·73 (0·72–0·74) | 0·86 (0·84–0·87) | | 0·91 (0·88–0·93) | –0·07 (–0·09––0·04) | |
| All ethnic groups | – | 0·6 (0·53–0·69) | – | – | | | – |

### Table S12: Effect modification by ethnicity for the effect of migration on pre–pandemic consultation rates (18–category ethnic groups)

|  | **RR (95% CI) comparing each group to White British non–migrants** | | **RR (95% CI) comparing migrants to non–migrants of each ethnicity** | | **Multiplicative effect (95%CI)** | **RERI, additive effect (95%CI)** | |
| --- | --- | --- | --- | --- | --- | --- | --- |
| Ethnicity | Non–migrants | Migrants |  |  |  |  |  |
| British | 1·0 | 0·95  (0·92–0·97) | 0·95  (0·92–0·97) | | – | – | |
| Irish | 0·96  (0·93–0·99) | 0·76  (0·71–0·81) | 0·79  (0·74–0·85) | | 0·84  (0·77–0·91) | –0·15  (–0·21––0·08) | |
| Gypsy or Irish Traveller | 1·22  (1·03–1·45) | 1·13  (0·97–1·31) | 0·92  (0·74–1·16) | | 0·98  (0·78–1·23) | –0·04  (–0·31–0·23) | |
| Other White | 0·91  (0·9–0·92) | 0·69  (0·68–0·69) | 0·75  (0·74–0·76) | | 0·8  (0·77–0·82) | –0·17  (–0·2––0·15) | |
| Mixed White and Black Caribbean | 1·09  (1·04–1·14) | 0·73  (0·66–0·82) | 0·68  (0·6–0·76) | | 0·72  (0·63–0·81) | –0·3  (–0·39––0·2) | |
| Mixed White and Black African | 0·99  (0·93–1·04) | 0·84  (0·8–0·88) | 0·85  (0·79–0·92) | | 0·9  (0·83–0·98) | –0·09  (–0·16––0·02) | |
| Mixed White and Asian | 0·9  (0·86–0·95) | 0·89  (0·84–0·93) | 0·99  (0·92–1·06) | | 1·04  (0·97–1·12) | 0·04  (–0·03–0·11) | |
| Other Mixed | 0·92  (0·88–0·95) | 0·88  (0·84–0·92) | 0·96  (0·91–1·01) | 1·01  (0·96–1·08) | | | 0·02  (–0·04–0·07) |
| Indian | 1·02  (0·99–1·04) | 1·14  (1·12–1·16) | 1·12  (1·1–1·15) | 1·19  (1·15–1·23) | | | 0·18  (0·14–0·22) |
| Pakistani | 1·15  (1·12–1·19) | 1·28  (1·25–1·3) | 1·11  (1·07–1·15) | 1·17  (1·12–1·22) | | | 0·18  (0·13–0·23) |
| Bangladeshi | 1·27  (1·21–1·33) | 1·29  (1·26–1·33) | 1·02  (0·97–1·08) | 1·08  (1·02–1·14) | | | 0·08  (0·01–0·15) |
| Chinese | 0·64  (0·61–0·67) | 0·7  (0·68–0·73) | 1·1  (1·04–1·16) | 1·16  (1·1–1·23) | | | 0·12  (0·08–0·16) |
| Other Asian | 0·88  (0·86–0·91) | 1·21  (1·19–1·23) | 1·37  (1·33–1·41) | 1·45  (1·39–1·51) | | | 0·38  (0·34–0·42) |
| African | 0·86  (0·84–0·87) | 1·02  (1–1·04) | 1·19  (1·16–1·22) | 1·26  (1·21–1·31) | | | 0·22  (0·18–0·25) |
| Caribbean | 0·92  (0·89–0·95) | 0·97  (0·93–1·01) | 1·05  (1–1·11) | 1·11  (1·05–1·18) | | | 0·1  (0·05–0·16) |
| Other Black | 0·91  (0·88–0·93) | 0·92  (0·89–0·95) | 1·01  (0·97–1·06) | 1·07  (1·02–1·13) | | | 0·07  (0·02–0·11) |
| Arab | 0·99  (0·86–1·14) | 0·97  (0·91–1·03) | 0·98  (0·84–1·14) | 1·04  (0·89–1·21) | | | 0·04  (–0·12–0·19) |
| Any other ethnic group | 0·89  (0·86–0·92) | 0·9  (0·89–0·92) | 1·02  (0·98–1·05) | 1·07  (1·03–1·12) | | | 0·07  (0·03–0·11) |
| Unknown | 0·85  (0·85–0·86) | 0·73  (0·72–0·74) | 0·86  (0·84–0·87) | 0·91  (0·88–0·93) | | | –0·07  (–0·09––0·04) |

### Table S13: Effect modification by ethnicity for the effect of migration on consultation rates pre vs during the pandemic

| **Ethnicity** | **Variable** | **Interpretation** | **RR (95%CI)** |
| --- | --- | --- | --- |
| White British (ethnicity reference group) | 2–way interaction (between migration status and pandemic) | Effect of being a migrant of X ethnicity vs· non–migrant of that same ethnicity on consultation rate during the pandemic restrictions compared to before the pandemic (i·e· the additional effect of the pandemic on the way ethnicity modifies the association between migration and consultation rate) | 0·69 (0·64–0·73) |
| White non–British | 2–way interaction (between migration status and pandemic)  +  3–way interaction (between migration status, ethnicity and pandemic) |  | 0·72 (0·68–0·77) |
| Mixed/Multiple ethnic groups |  |  | 1·04 (0·95–1·14) |
| Asian/Asian British |  |  | 1·11 (1·04–1·18) |
| Black/African/ Caribbean/Black British |  |  | 0·68 (0·64–0·73) |
| Other ethnic group |  |  | 0·83 (0·75–0·91) |
| Unknown |  |  | 0·8 (0·75–0·86) |

### **Figure S2: Weekly consultation rates by migration status and ethnicity (18 categories) in England: predicted rates from interrupted time–series analysis (solid line) and actual observed rates (dashed line), truncated view July 2019–November 2020**

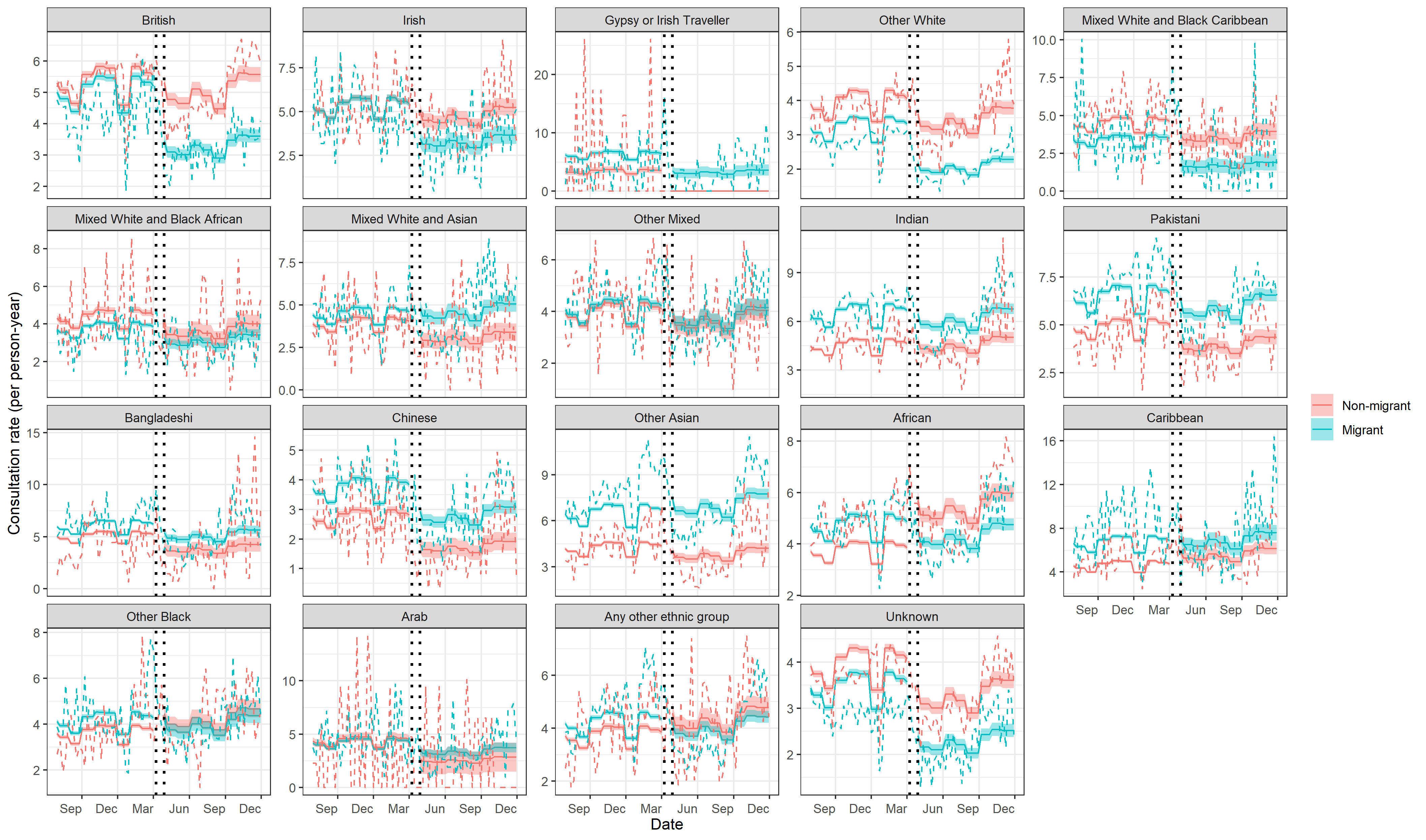

### Table S14: Effect modification by ethnicity (18 categories) for the effect of migration on consultation rates pre vs during the pandemic

| **Ethnicity** | **Variable** | **Interpretation** | **RR (95%CI)** |
| --- | --- | --- | --- |
| British (ethnicity reference group) | 2–way interaction (between migration status and pandemic) | Effect of being a migrant of X ethnicity vs· non–migrant of that same ethnicity on consultation rate during the pandemic restrictions compared to before the pandemic (i·e· the additional effect of the pandemic on the way ethnicity modifies the association between migration and consultation rate) | 0·69 (0·64–0·74) |
| Irish | 2–way interaction (between migration status and pandemic)  +  3–way interaction (between migration status, ethnicity and pandemic) |  | 0·7 (0·59–0·83) |
| Gypsy or Irish Traveller |  |  | –* |
| Other White |  |  | 0·74 (0·69–0·79) |
| Mixed White and Black Caribbean |  |  | 0·63 (0·46–0·87) |
| Mixed White and Black African |  |  | 0·99 (0·85–1·16) |
| Mixed White and Asian |  |  | 1·31 (1·1–1·57) |
| Other Mixed |  |  | 0·99 (0·87–1·13) |
| Indian |  |  | 0·94 (0·86–1·02) |
| Pakistani |  |  | 1·13 (1·02–1·25) |
| Bangladeshi |  |  | 1·12 (0·94–1·33) |
| Chinese |  |  | 1·18 (0·99–1·41) |
| Other Asian |  |  | 1·2 (1·09–1·32) |
| African |  |  | 0·63 (0·59–0·68) |
| Caribbean |  |  | 0·85 (0·75–0·97) |
| Other Black |  |  | 0·82 (0·73–0·92) |
| Arab |  |  | 1·37 (0·83–2·27) |
| Any other ethnic group |  |  | 0·82 (0·74–0·91) |
| Unknown |  |  | 0·8 (0·74–0·86) |

*RR not available due to small numbers in the group

### Figure S3: Forest plot of pre–pandemic sensitivity analyses in England: consultation rate ratios comparing migrants and non–migrants matched by (1) age and year of joining the CPRD GOLD database and practice region and (2) by follow–up time in person–years and practice region

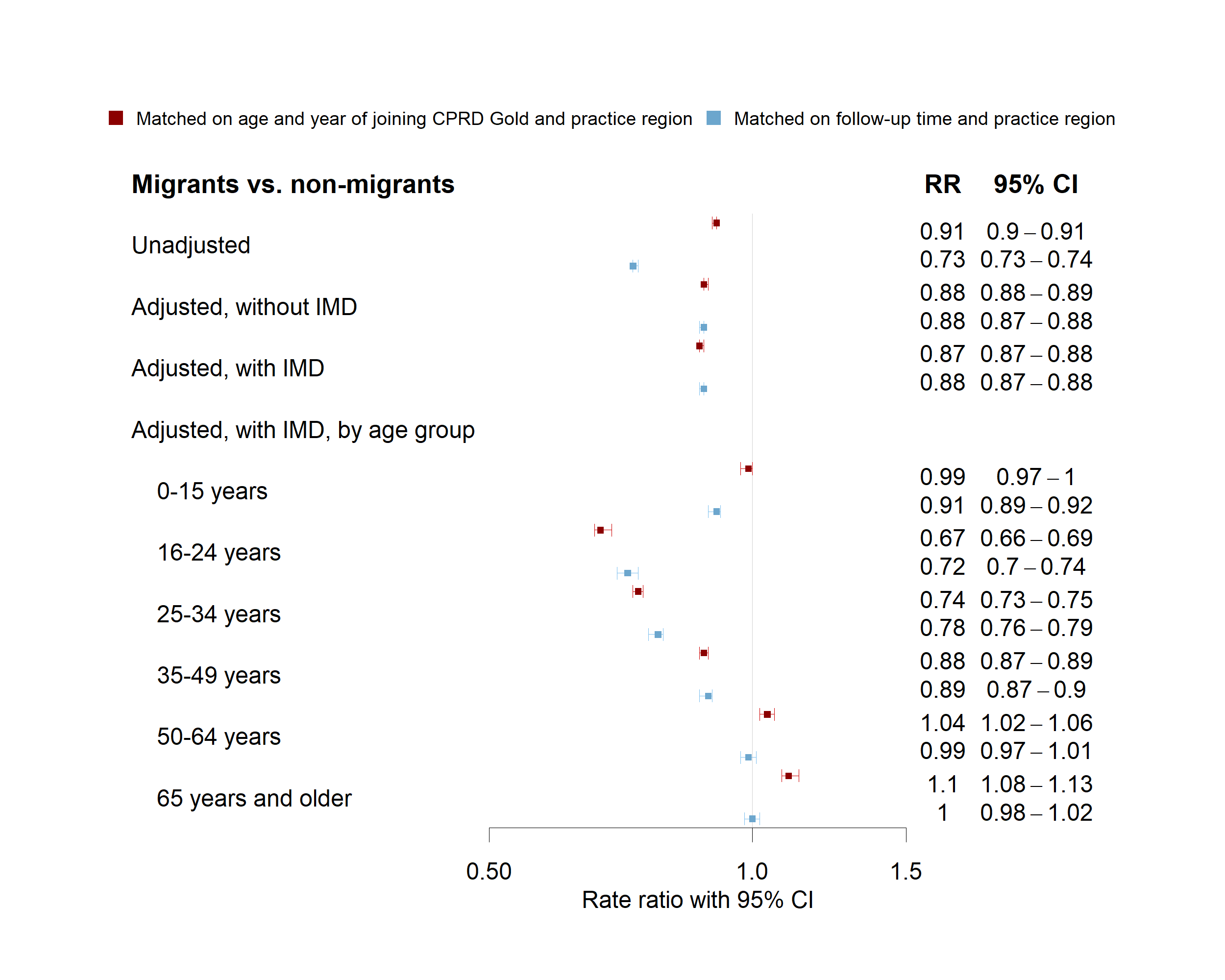

### Figure S4: Forest plot of pre–pandemic sensitivity analyses in England: consultation rate ratios by migration certainty

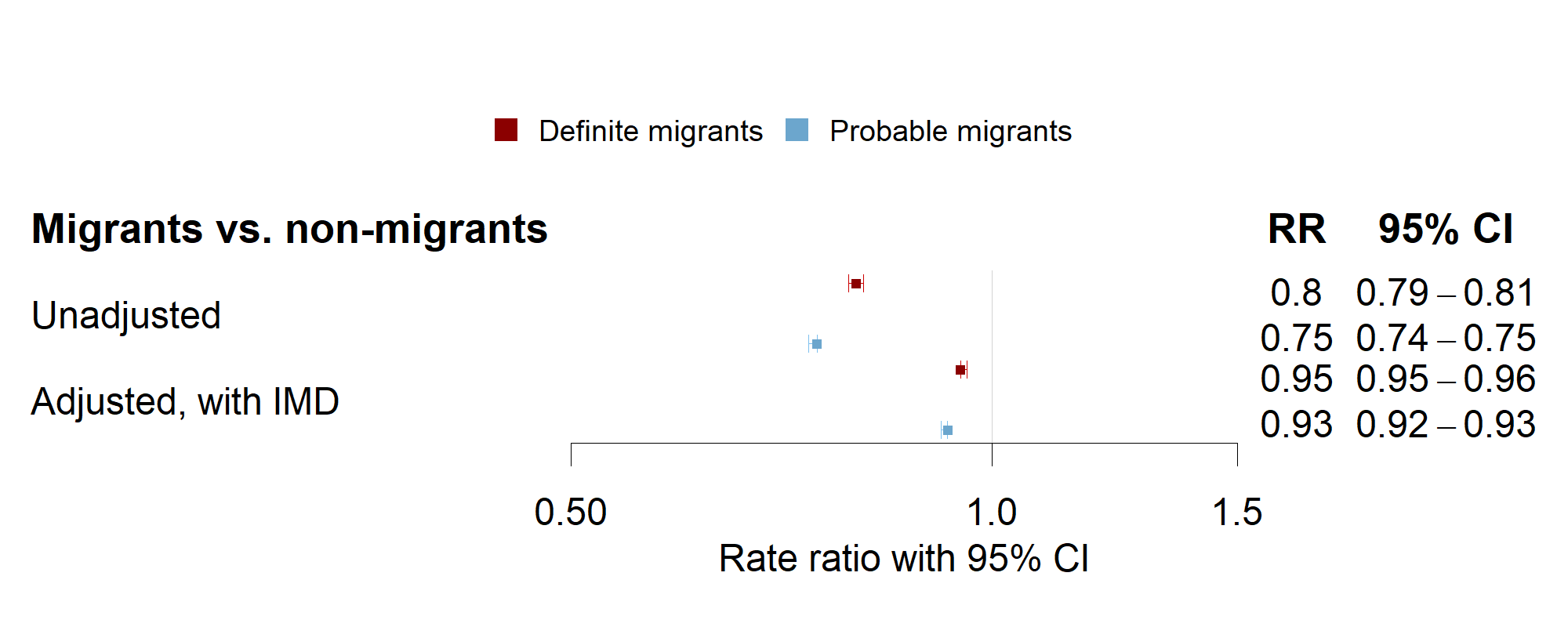

### Figure S5: Weekly consultation rates by migration certainty in England: predicted rates from interrupted time–series analysis (solid line) and actual observed rates (dashed line), truncated view July 2019 – November 2020

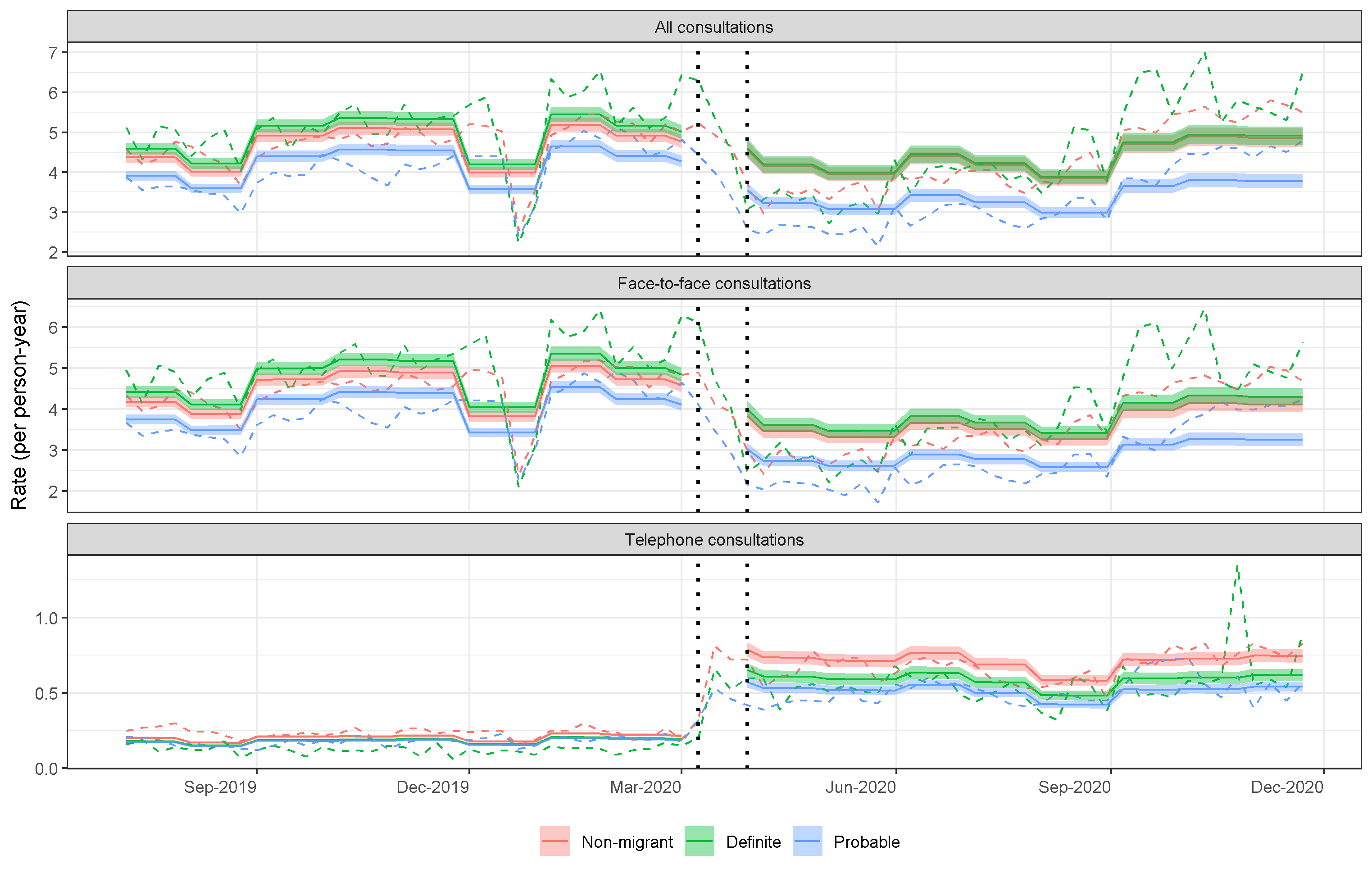

### Table S15: Consultation rate ratios from interrupted time series sensitivity analysis for England by migration certainty (5 January 2015 to 26 December 2020)

| Variable | Interpretation | All | Face–to–face | Telephone |
| --- | --- | --- | --- | --- |
| Pandemic | Comparing non–migrants’ consultation rates during the pandemic to non–migrants’ consultation rates pre–pandemic | 0·96 (0·91–1) | 0·84 (0·8–0·87) | 3·7 (3·5–3·92) |
| ‘Definite’ migrant | Comparing ‘definite’ migrants’ consultation rates pre–pandemic to non–migrants consultations pre–pandemic | 1·05 (1·03–1·07) | 1·06 (1·04–1·08) | 0·89 (0·87–0·92) |
| ‘Probable’ migrant | Comparing ‘probable’ migrants’ consultation rates pre–pandemic to non–migrants consultations pre–pandemic | 0·9 (0·88–0·91) | 0·9 (0·88–0·92) | 0·87 (0·85–0·89) |
| ‘Definite’ migrant  +  ‘Definite’ migrant interaction | Comparing ‘definite’ migrants’ consultation rates during the pandemic to non–migrants’ consultation rates during the pandemic | 1·01 (0·95–1·07) | 1·05 (0·99–1·11) | 0·83 (0·77–0·89) |
| ‘Probable’ migrant  +  ‘Probable’ migrant interaction | Comparing ‘probable’ migrants’ consultation rates during the pandemic to non–migrants’ consultation rates during the pandemic | 0·78 (0·73–0·82) | 0·79 (0·75–0·83) | 0·73 (0·68–0·78) |
| ‘Definite’ migrant interaction (between migration certainty and pandemic) | Comparing the difference in consultation rates between ‘definite’ migrants and non–migrants pre–pandemic to the difference in consultation rates between ‘definite’ migrants and non–migrants during the pandemic (the additional effect of the pandemic on the difference between ‘definite’ migrants and non–migrants i·e· the multiplicative effect) | 0·96 (0·91–1·02) | 0·99 (0·93–1·05) | 0·93 (0·86–1·01) |
| ‘Probable’ migrant interaction (between migration certainty and pandemic) | Comparing the difference in consultation rates between ‘probable’ migrants and non–migrants pre–pandemic to the difference in consultation rates between ‘probable’ migrants and non–migrants during the pandemic (the additional effect of the pandemic on the difference between ‘probable’ migrants and non–migrants i.e. the multiplicative effect) | 0·87 (0·82–0·92) | 0·88 (0·83–0·93) | 0·83 (0·77–0·9) |

### Figure S6: Weekly consultation rates by migration status in England: predicted rates (solid line) from interrupted time series sensitivity analysis (matched on age and year of joining the CPRD GOLD database, sex, practice region and IMD) and observed rates (dashed line), truncated view July 2019 – November 2020

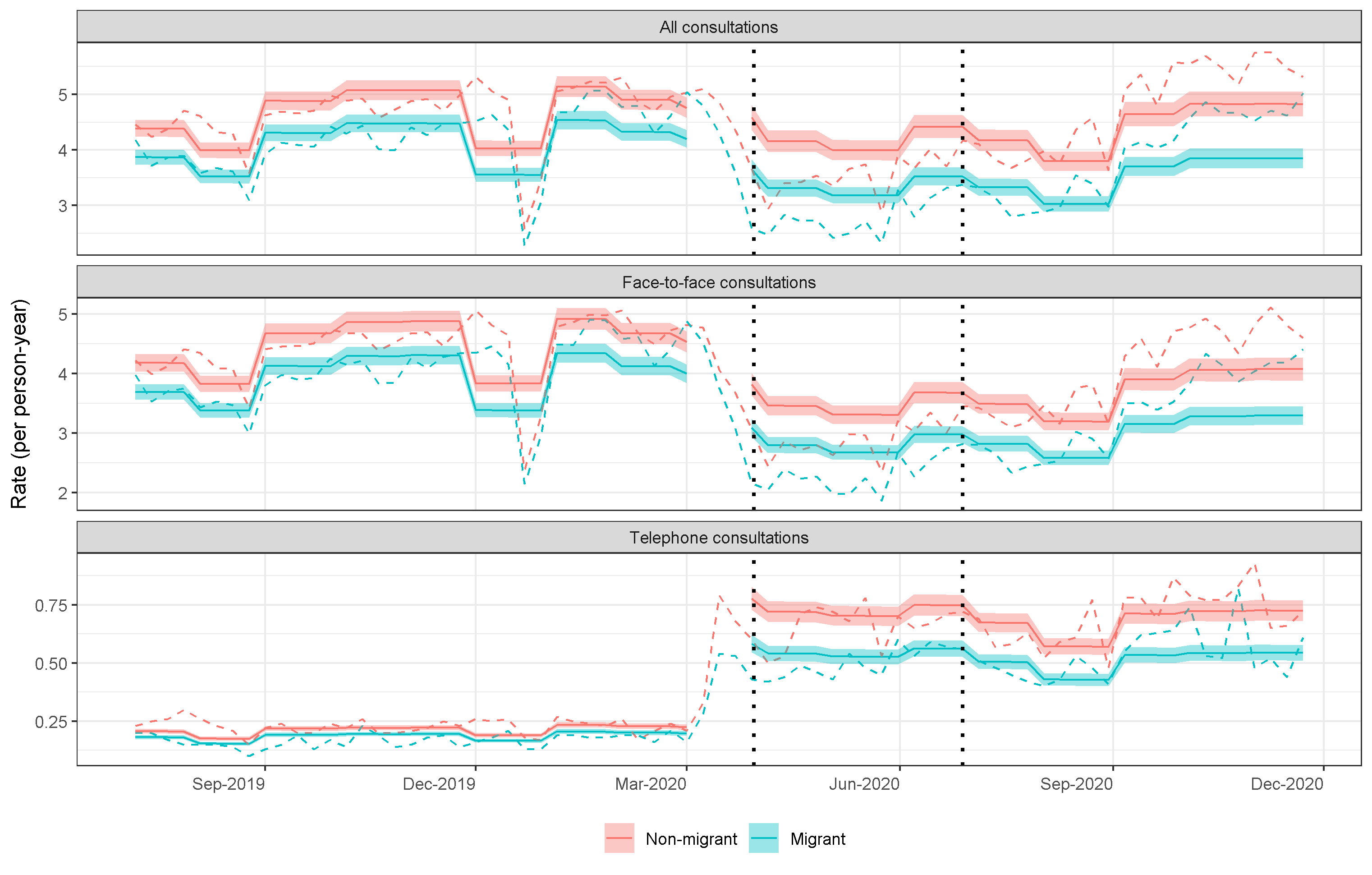

### Table S16: Consultation rate ratios from interrupted time series analysis (5 January 2015 to 26 December 2020) for England in cohort matched on age and year of joining the CPRD GOLD database, sex, practice region and IMD

| Variable | Interpretation | All | Face–to–face | Telephone |
| --- | --- | --- | --- | --- |
| Pandemic | Comparing non–migrants’ consultation rates during the pandemic to non–migrants’ consultation rates pre–pandemic | 0·96  (0·92–1·01) | 0·84  (0·81–0·88) | 3·46  (3·27–3·67) |
| Migration status | Comparing migrants’ consultation rates pre–pandemic to non–migrants consultations pre–pandemic | 0·88  (0·87–0·9) | 0·88  (0·87–0·9) | 0·88  (0·86–0·9) |
| Migration status +  interaction term (between migrant status and pandemic) | Comparing migrants’ consultation rates during the pandemic to non–migrants’ consultation rates during the pandemic | 0·8  (0·76–0·84) | 0·81  (0·77–0·85) | 0·75  (0·7–0·8) |
| Interaction term (between migrant status and pandemic) | Comparing the difference in consultation rates between migrants and non–migrants pre–pandemic to the difference in consultation rates between migrants and non–migrants during the pandemic (the additional effect of the pandemic on the difference between migrants and non–migrants i.e. the multiplicative effect) | 0·9  (0·86–0·95) | 0·92  (0·87–0·97) | 0·85  (0·8–0·92) |

### STROBE checklist

|  | Item No. | | Recommendation | | | Page  No. |
| --- | --- | --- | --- | --- | --- | --- |
| **Title and abstract** | 1 | | (*a*) Indicate the study’s design with a commonly used term in the title or the abstract | | | Title – p1 |
|  |  |  | (*b*) Provide in the abstract an informative and balanced summary of what was done and what was found | | | Abstract – p2 |
| Introduction | | | | | | |
| Background/rationale | 2 | | Explain the scientific background and rationale for the investigation being reported | | | Background – p4 |
| Objectives | 3 | | State specific objectives, including any prespecified hypotheses | | | Background – p4 |
| Methods | | | | | | |
| Study design | 4 | | Present key elements of study design early in the paper | | | Title – p1  Abstract – p2  Methods/Study design and data management – p4 |
| Setting | 5 | | Describe the setting, locations, and relevant dates, including periods of recruitment, exposure, follow-up, and data collection | | | Methods/Study design and data management – p4  Methods/Study cohort – p4/5 |
| Participants | 6 | | (*a*) *Cohort study*—Give the eligibility criteria, and the sources and methods of selection of participants. Describe methods of follow-up  *Case-control study*—Give the eligibility criteria, and the sources and methods of case ascertainment and control selection. Give the rationale for the choice of cases and controls  *Cross-sectional study*—Give the eligibility criteria, and the sources and methods of selection of participants | | | Methods/Study design and data management – p4  Methods/Study cohort – p4/5  Supplementary Information/Supplementary Box 1 – p3 |
|  |  |  | (*b*) *Cohort study*—For matched studies, give matching criteria and number of exposed and unexposed  *Case-control study*—For matched studies, give matching criteria and the number of controls per case | | |  |
| Variables | 7 | | Clearly define all outcomes, exposures, predictors, potential confounders, and effect modifiers. Give diagnostic criteria, if applicable | | | Methods/Exposure and outcomes – p4  Methods/Statistical analysis: Before the pandemic – p7  Methods: Statistical analysis: Effect modification by ethnicity – p7 |
| Data sources/ measurement | 8* | | For each variable of interest, give sources of data and details of methods of assessment (measurement). Describe comparability of assessment methods if there is more than one group | | | Methods/Study design and data management – p4  Methods/Exposure and outcomes – p4  Methods/Figure 1 – p6 |
| Bias | 9 | | Describe any efforts to address potential sources of bias | | | Methods/Bias – p7 |
| Study size | 10 | | Explain how the study size was arrived at | | | Methods/Study cohort – p4/5  Methods/Figure 1 – p6 |
| Quantitative variables | 11 | | Explain how quantitative variables were handled in the analyses. If applicable, describe which groupings were chosen and why | | | Methods/Exposure and outcomes – p4  Methods/Study cohort – p4/5  Information/Supplementary Box 1 – p3 |
| Statistical methods | 12 | | (a) Describe all statistical methods, including those used to control for confounding | | | Methods/Statistical analysis: Before the pandemic – p7  Methods/Statistical analysis: Before versus during the pandemic – p7  Methods: Statistical analysis: Effect modification by ethnicity – p7  Supplementary Box 2: Interrupted time-series statistical analysis – p4 |
|  |  | | (b) Describe any methods used to examine subgroups and interactions | | | Methods: Statistical analysis: Effect modification by ethnicity – p7  Supplementary Information/Supplementary Box 2: Interrupted time-series statistical analysis – p4 |
|  |  | | (c) Explain how missing data were addressed | | | N/A for main migration exposure  Methods: Statistical analysis: Effect modification by ethnicity – p7 (for missing ethnicity data) |
|  |  | | (d) Cohort study—If applicable, explain how loss to follow-up was addressed  Case-control study—If applicable, explain how matching of cases and controls was addressed  Cross-sectional study—If applicable, describe analytical methods taking account of sampling strategy | | | N/A |
|  |  | | (e) Describe any sensitivity analyses | | | Methods/Bias – p7 |
| Results | | | |  |  | |
| Participants | 13* | (a) Report numbers of individuals at each stage of study—e.g. numbers potentially eligible, examined for eligibility, confirmed eligible, included in the study, completing follow-up, and analysed | | | | Methods/Figure 1 – p6 |
|  |  | (b) Give reasons for non-participation at each stage | | | |  |
|  |  | (c) Consider use of a flow diagram | | | |  |
| Descriptive data | 14* | (a) Give characteristics of study participants (e.g. demographic, clinical, social) and information on exposures and potential confounders | | | | Results/Table 1 – p9  Supplementary Information/Table S3 – 12  Supplementary Information/Table S4 – 14  Supplementary Information/Table S5 – 16  Supplementary Information/Table S6 – 18 |
|  |  | (b) Indicate number of participants with missing data for each variable of interest | | | | Missing ethnicity data:  Results/Table 1 – p9  Supplementary Information/Table S3 – 12  Supplementary Information/Table S4 – 14  Supplementary Information/Table S5 – 16  Supplementary Information/Table S6 – 18 |
|  |  | (c) *Cohort study*—Summarise follow-up time (e.g., average and total amount) | | | | Results/Table 1 – p9  Supplementary Information/Table S3 – 12  Supplementary Information/Table S4 – 14  Supplementary Information/Table S5 – 16  Supplementary Information/Table S6 – 18 |
| Outcome data | 15* | *Cohort study*—Report numbers of outcome events or summary measures over time | | | | Supplementary Information/Table S7 – p20  Supplementary Information/Table S8 – p21 |
|  |  | *Case-control study—*Report numbers in each exposure category, or summary measures of exposure | | | | N/A |
|  |  | *Cross-sectional study—*Report numbers of outcome events or summary measures | | | | N/A |
| Main results | 16 | (*a*) Give unadjusted estimates and, if applicable, confounder-adjusted estimates and their precision (e.g., 95% confidence interval). Make clear which confounders were adjusted for and why they were included | | | | Results/Before the pandemic – p11  Results/Figure 2 – p12  Results/Before versus during the pandemic – p12/13  Results/Figure 3 – p13  Results/Table2 – p13/14 |
|  |  | (*b*) Report category boundaries when continuous variables were categorized | | | | N/A |
|  |  | (*c*) If relevant, consider translating estimates of relative risk into absolute risk for a meaningful time period | | | | N/A |
| Other analyses | 17 | Report other analyses done—e.g. analyses of subgroups and interactions, and sensitivity analyses | | | | Results/Figure 2B – p12  Results/Figure 4 – p14  Results/Figure 5 – p15  Supplementary Information/Tables S9 – S16 – p22, 25–27, 29, 33, 35  Supplementary Information/Figures S1–S6 – p23, 28, 30–32, 34 |
| **Discussion** |  |  | | | |  |
| Key results | 18 | Summarise key results with reference to study objectives | | | | Discussion – p17 |
| Limitations | 19 | Discuss limitations of the study, taking into account sources of potential bias or imprecision. Discuss both direction and magnitude of any potential bias | | | | Discussion – p17 |
| Interpretation | 20 | Give a cautious overall interpretation of results considering objectives, limitations, multiplicity of analyses, results from similar studies, and other relevant evidence | | | | Discussion – p17/18 |
| Generalisability | 21 | Discuss the generalisability (external validity) of the study results | | | | Discussion – p18 |
| **Other information** |  |  | | | |  |
| Funding | 22 | Give the source of funding and the role of the funders for the present study and, if applicable, for the original study on which the present article is based | | | | Methods/Role of the funding source – p7  Additional Information/Funding – p19 |
